# Unraveling the genetic architecture of the human mammillary bodies

**DOI:** 10.64898/2026.11.09.26367603

**Authors:** Mohammed Aslam Imtiaz, Peng Xu, Santiago Estrada, Mohamed M. Saleh, Isaac S. Racine, Dan Liu, Monique M.B. Breteler, N. Ahmad Aziz

## Abstract

The mammillary bodies are a central component of the hippocampal–diencephalic-cingulate network that is critically involved in emotion processing and memory function, yet their genetic architecture remains unknown. We conducted a genome-wide association study (GWAS) of mammillary body volume in 38,064 participants from the UK Biobank and replicated our findings in the Rhineland Study cohort (N = 4,860). Mammillary body and hypothalamic subregional volumes were derived from structural brain MRIs using FastSurfer-HypVINN, an established and validated deep learning-based segmentation tool. Univariate GWAS identified 34 genomic loci associated with mammillary body volume, including 26 novel loci, with 20 loci showing replication in the Rhineland Study. Meta-analysis across both cohorts (N = 42,924) identified 44 associated loci, of which 35 were novel. Multivariate GWAS of hypothalamic subregions identified 55 loci, including 40 novel associations. Integrative cross-omics analyses identified *MEI1* and CAPZA1 as key molecular candidates, with genetically regulated *MEI1* expression and circulating CAPZA1 protein levels partially mediating genetic effects on mammillary body volume. Genetic correlation and Mendelian randomization analyses further revealed a bidirectional relationship between mammillary body volume and major depressive disorder. Together, our findings elucidate the genetic architecture of mammillary body volume and its relation to mental disorders.

## INTRODUCTION

The mammillary bodies are a pair of small, rounded nuclei located on the posteroinferior surface of the hypothalamus (1). They are a central component of the Papez circuit (2), receiving input from the hippocampus via the fornix and projecting predominantly through the mammillothalamic tract to the anterior thalamic nuclei. This network plays a fundamental role in emotion and memory processing, particularly episodic and spatial memory consolidation (1). In addition to hippocampal inputs, the mammillary bodies also receive inputs from brainstem tegmental nuclei (1), indicating a broader role in limbic circuitry, beyond a simple hippocampal relay function.

The mammillary bodies have been implicated in different neurological and psychiatric diseases, the most prominent being Wernicke-Korsakoff syndrome, in which mammillary body dysfunction leads to severe anterograde amnesia (3). Furthermore, postmortem studies have found alterations in mammillary body volume in patients with bipolar disorder and a significant volume reduction in major depressive disorder (MDD) compared to individuals without heart failure (5), suggesting an unrecognized role in these two common affective disorders as well. Another postmortem study showed selective vulnerability and neuronal degeneration of mammillary body neurons in Alzheimer’s disease, suggesting that dysfunction of this structure may also contribute neurodegeneration (3). In addition, in an analysis of two large population-based cohorts, we recently demonstrated that mammillary body volume substantially decreases with age in both sexes (4). Conversely, larger mammillary body volume was associated with better global cognition as well as better domain-specific cognitive performance, including working memory, episodic memory, executive function and processing speed (4). Together, these findings support the relevance of the mammillary bodies in affective diseases and age-associated cognitive decline, warranting further scrutiny of the determinants of their structure and function.

Surprisingly, despite their critical role in cognitive and affective processes, the genetic and molecular mechanisms underlying interindividual differences in mammillary body volume remain largely unknown. Although a recent genome-wide association study (GWAS) of the hypothalamus and its subregions identified 23 genomic loci associated with the overall volume of the hypothalamus and its neighboring subregions, the mammillary bodies were analyzed only as part of the posterior hypothalamus, together with the lateral hypothalamus and the tuberomammillary nucleus (5). However, the genetic architecture specific to the mammillary bodies could not be resolved independently from these neighboring structures. Moreover, brain images were analyzed using an earlier hypothalamic segmentation tool (6), which was trained on manually labeled hypothalamic subunits derived from T1-weighted MRI scans. More advanced, multimodal, fully automated segmentation tools of the hypothalamus and its adjacent structures have been developed since (7), which enable a more fine-grained delineation of hypothalamic substructures, including distinct parcellation of the mammillary bodies.

Using the FastSurfer-HypVINN tool, which enables accurate multimodal segmentation of the hypothalamus and its subregions, we performed a GWAS of mammillary body volume, followed by a multivariate GWAS incorporating all major hypothalamic subregions, including the anterior, medial, tuberal, lateral and posterior subregions. We estimated the heritability of mammillary body volume and other hypothalamic subregions using linkage disequilibrium score regression (LDSC) and assessed the genetic overlap with volumetric measures in other brain regions. In addition, to investigate the relationships with mood-related traits, psychiatric disorders, and neurodegenerative diseases, we examined genetic correlations between these outcomes and mammillary body volume and conducted causal inference analyses using two-sample Mendelian randomization (MR) analyses. For the genomic loci identified to be associated with mammillary body volume, we also performed expression quantitative trait loci (eQTL) and protein quantitative trait loci (pQTL) analyses. Lastly, we evaluated gene and protein expression associations with mammillary body volume. Overall, through this integrative, multi-omics approach we identified several key genes and proteins that underpin the structural integrity of the mammillary bodies and found (potentially causal) relationships between mammillary body volume and psychiatric, neurodevelopmental and neuro-degenerative disorders.

## METHODS

### Study population

For the discovery GWAS we included 38,064 participants from the UK Biobank (UKBB) with available structural brain MRI scans and genetic data. For replication analyses, we included 4,860 participants from the Rhineland Study, who had structural brain MRI scans and genetic data available. Imaging datasets that failed quality control following visual inspection of the MRI scans were excluded from the analyses. By combining the discovery and replication cohort, we performed a GWAS meta-analysis with a total sample size of 42,924 participants. The UKBB study was approved by the National Information Governance Board for Health and Social Care and the National Health Service North West Centre for Research Ethics Committee (Ref: 11/NW/0382). The Rhineland Study was approved by the Ethics Committee of the Medical Faculty of the University of Bonn and conducted in accordance with the Declaration of Helsinki. All participants provided written informed consent.

### Imaging data

In UKBB, brain imaging data were acquired using Siemens Skyra 3T MRI scanners equipped with 32-channel head coils. Imaging was initially conducted at a single scanning site in Cheadle (Manchester, UK) using one scanner and was later expanded to include two additional scanners located in Newcastle and Reading (8). Detailed information on the MRI acquisition protocol and sequence parameters is available on the UKBB website (biobank.ctsu.ox.ac.uk/crystal/crystal/docs/brain_mri.pdf). In the Rhineland Study, imaging data were acquired using two identical 3T Siemens MAGNETOM Prisma MRI scanners equipped with 64-channel head–neck coils. MRI scans were acquired at two examination centers in Bonn, Germany (9).

Segmentation of the mammillary bodies and hypothalamic subregions, including the anterior, medial, lateral, tuberal, and posterior region, was performed using the fully automated deep learning–based segmentation tool FastSurfer-HypVINN (7). In the Rhineland Study, segmentation was performed using 0.8-mm isotropic T1- and T2-weighted images, whereas in UKBB, 1.0-mm isotropic T1-weighted images were used. FastSurfer-HypVINN has previously been validated against manually segmented ground-truth MRI data from both the UKBB and the Rhineland Study. Estimated total intracranial volume (eTIV) was derived for both cohorts using the standard FreeSurfer (version 6.0) processing pipeline (10).

For image quality control within the Rhineland Study, all scans underwent visual inspection based on predefined criteria. Conversely, due to the larger sample size of the UKBB Imaging Study, we implemented a targeted approach: 1,561 scans were initially flagged as statistical outliers because their volumetric measures deviated by more than 2 standard deviations from the mean. All of these flagged datasets were subsequently reviewed manually. Additionally, a random subset of 500 scans from the remaining pool was visually inspected to evaluate segmentation quality, yielding a 96.4% pass rate. Exclusion criteria included severe motion artifacts, poor tissue contract, or incomplete hypothalamic subregions coverage that compromised reliable segmentation.

### Genotyping data

In UKBB, genetic quality control was performed according to standard protocols (11). Single nucleotide polymorphisms (SNPs) were excluded based on low call rate (<95%), minor allele frequency (MAF<1%), and deviation from Hardy–Weinberg equilibrium (HWE; p < 1×10). At the sample level, participants were excluded based on low call rate, abnormal heterozygosity, and non-European ancestry. Genotype imputation was performed using the Haplotype Reference Consortium (HRC) imputation panel. In the Rhineland Study, genotyping was performed using the Illumina Omni 2.5 Exome Array. Signal intensity files were processed using GenomeStudio (version 2.0.5). SNP and sample-level quality control procedures were conducted using PLINK (version 1.9). SNPs were excluded based on missingness (>2%), deviation from HWE (p < 1×10), and MAF <1%. Samples were excluded based on low call rate, abnormal heterozygosity, and non-European ancestry, which was determined using principal component analysis implemented in EIGENSTRAT (version 6.1.4) (12). Quality-controlled genotypes were imputed to the HRC reference panel (version r1.1) using the Helmholtz Munich Imputation Server (13).

### Transcriptomic analysis

We functionally validated our results using gene expression data of the first 3000 consecutively enrolled participants of the Rhineland Study. Samples used for RNA sequencing were stored in PAXgene Blood RNA tubes (PreAnalytix/Qiagen), and were thawed and incubated at room temperature to increase RNA yields. Total RNA was isolated according to the manufacturer’s instructions using PAXgene Blood miRNA Kit following their automated purification protocol (PreAnalytix/Qiagen). RNA integrity and quantity were evaluated using the TapeStation RNA assay on a TapeStation4200 instrument (Agilent). The Tapestation RNA assay (Tapestation4200 instrument from Agilent). After using 750 ng of total RNA to generate next generation-sequencing libraries for total RNA sequencing (TruSeq stranded total RNA kit, Illumina), a Ribo-Zero Globin reduction was performed. Libraries were quantified using Qubit HS dsDNA assay (Invitrogen) and clustered at 250 pM concentrations on a NovaSeq6000 instrument. Quality control of the sequencing reads was performed with FastQC (v0.11.9). Following the filtering of low-quality score reads with Trimmomatic (v.0.39), sequencing reads were aligned to the human reference genome GRCh38.p13 using STAR (v2.7.1). The count matrix was generated with STAR –quantMode GeneCounts using the human gene annotation version GRCh38.101. Genes with overall mean expression > 15 reads and expressed in at least 5% of the participants were retained for further analysis. Raw counts were normalized and transformed using the varianceStabilizingTransformation function from DESeq2 (v1.30.1). Following quality control and merging with genomic and imaging data, 1,891 participants were included in the downstream eQTL and gene-expression association analyses.

### Proteomic analysis using OLINK

In the Rhineland Study, among the first 5,500 participants enrolled before April 2020, 2,852 were selected using age- and sex-stratified random sampling for proteomics analysis. Plasma proteins were measured using the Olink Explore platform, which incorporates internal and external quality control (QC) procedures to ensure assay and sample reliability. Two QC metrics were evaluated: QC_warning at the sample level and Assay_warning at the protein level. Samples received a QC_warning (WARN) if average matched counts were <500 or if Incubation or Amplification Control deviated by ±0.3 NPX from the plate median. Manually flagged samples were labeled MANUAL_WARN. At the assay level, an Assay_warning (WARN) was assigned if the median of triplicate negative controls deviated by >5 SD from predefined validation thresholds. As an initial filtering step, proteins flagged as “Excluded” were removed; if a protein was excluded within a sample-panel combination, all proteins from that panel in that sample were removed. In total, 56 proteins were excluded. Outlier detection was performed within each panel using: 1) principle component analysis (PC1 or PC2 > ±6 SD), and 2) NPX distribution metrics (median or interquartile range > ±6 SD). One sample was consistently identified as an outlier across all panels and was excluded. In the final dataset comprising 2,888 protein assays, 123 assays (4.3%) had Assay_warning flags. QC_warning percentages ranged from 0% to 8.6%, with higher rates mainly in the Oncology II panel. Most assays showed 0% warnings, indicating high technical reliability. Assay warnings ≥ 5% were rare and primarily observed in the Oncology II and Inflammation II panels. Following quality control and merging with genomic and imaging data, 2,299 participants were included in the downstream OLINK-based pQTL and protein association analyses.

### Proteomic analysis using SEER

Among participants enrolled in the Rhineland Study before November 26, 2021, 551 were selected randomly after stratifying by age and sex, creating a random representative subset of the entire cohort. Plasma (240 μl) from each participant was sent to Seer Inc.® for quantification with the Proteograph XT platform, providing untargeted and deep proteomic profiling via liquid chromatography-mass spectrometry. Liquid chromatography was performed via the Thermo Scientific Vanquish Neo UHPLC system, while mass spectrometry was performed via two Thermo Fisher Orbitrap Astral instruments. Peptide identification relied on the data-independent acquisition workflow, DIA-NN, using search engine v1.8.1 with Seer’s in-house database, uniprot_homosapiens_isoforms_con_20221209.fasta. Additionally, library-free *in silico* searches were performed using the UniProt reference database. Seer applied a 1% false discovery rate threshold for the identification of peptide and protein group levels. An updated internal Seer pipeline was used for additional quality control. The pipeline first median-normalized the data within each LC-MS run, using precursors detected in 80% or more of the samples. The MaxLFO algorithm was then used to combine the separate XT nanoparticle wells into a single protein abundance estimate for each sample. Finally, plate-level batch effects were corrected using the LIMMA R. This returned 7,960 uniquely identified proteins across the 551 samples. Protein intensities were then rank-transformed and principle component analysis was run on these transformed intensities. Only proteins with intensity measurements available in at least 10% of samples were included in downstream analyses. Following quality control and merging with genomic and imaging data, 376 participants were included in the downstream SEER-based pQTL and protein association analyses.

### Univariate GWAS

Univariate GWASs were conducted separately for each phenotype including mammillary body volume and volumes of all other hypothalamic subregions using the univariate framework implemented in MOSTest (14). For each phenotype, residuals were calculated after adjusting for age, age², sex, top 10 genetic principal components, eTIV, and cortical volumes of the frontal, parietal, temporal, and occipital lobes. In addition, analyses accounted for cohort-specific technical covariates, including MRI scanner site, batch effects, and genotyping array where applicable. Rank-based inverse normal transformation was applied to each phenotype prior to the MOSTest (14) analysis. SNP-wise Z-statistics were obtained for each phenotype from the univariate GWAS.

### Multivariate GWAS

The univariate Z-scores were subsequently used as inputs for the multivariate analysis. The phenotypic covariance structure was estimated using genome-wide Z-scores under permutation to model the null distribution while accounting for correlations among traits using MOSTest (14). For each SNP, a multivariate test statistic was computed that integrates the vector of univariate GWAS Z-scores while adjusting for their covariance structure, thereby capturing distributed genetic effects across correlated hypothalamic subregions. Under the null hypothesis of no association, the test statistic follows a chi-square distribution with degrees of freedom equal to the number of phenotypes. MOSTest fits a gamma distribution to the empirical null obtained from permutations to accurately estimate tail probabilities and extrapolate genome-wide significant p-values below 5 × 10□□ without requiring an infeasible number of permutations. Genome-wide significance was defined at p < 5 × 10□□.

### Meta-analysis GWAS

The univariate GWAS summary statistics for mammillary body volume were used to perform a GWAS meta-analysis using METAL (15). We performed a fixed-effects, inverse variance-weighted meta-analysis using the GWAS summary statistics from both cohorts.

### Definition of genomic risk loci

The FUMA (16) platform was used to identify genomic risk loci for univariate and multivariate GWAS. Genomic risk loci were defined as genomic regions containing independent significant SNPs and the SNPs in high linkage disequilibrium (r² > 0.6) with them. LD blocks of independent significant SNPs located within 250 kb of each other were merged into a single genomic risk locus. Lead SNPs were defined as independent significant SNPs with r² < 0.1 with one another. We used the 1000 Genomes Phase 3 (version 5) reference panel (17) for LD calculation and loci mapping.

### Gene and functional mapping

Using ANNOVAR (18), SNPs were positionally mapped to genes located within 10 kb of the variant. Using eQTL mapping, SNPs were further mapped to genes expressed in brain tissue databases, including GTEx v8 (19), BRAINEAC (20), PsychENCODE (21), and the CommonMind Consortium (22). Using chromatin interaction mapping, genes involved in significant chromatin interactions with SNP-associated regions were identified using datasets from PsychENCODE EP links (20), PsychENCODE promoter-anchored loops (22), Hi-C data from brain tissues (including adult cortex (23), fetal cortex (23), dorsolateral prefrontal cortex (24), and hippocampus (24)), Hi-C data from neural progenitor cell lines (24), and enhancer annotations from 17 brain-related repositories of the Roadmap Epigenomics Project (25) (i.e., E053, E054, E067, E068, E069, E070, E071, E072, E073, E074, E081, E082, E003, E008, E007, E009, E010, E057). Enhancer regions were defined as 250 bp upstream and 500 bp downstream of transcription start sites. Interactions passing the default false discovery rate (FDR) threshold (p < 1 × 10□□) were visualized using Circos plots. Using the GENE2FUNC tool within FUMA, we further investigated the biological context of the mapped genes by examining their enrichment in 53 GTEx tissue-specific gene expression sets (19), curated MSigDB gene sets (26), and the GWAS Catalog (27) of previously reported genetic variant–trait associations. Gene sets were considered significant if at least two input genes overlapped with the tested gene set and survived Bonferroni-adjusted p < 0.05.

### Gene-based association and gene-set enrichment analyses

Gene-based analyses were conducted using GWAS summary statistics from the univariate, multivariate, and meta-GWAS analyses in MAGMA (28). For gene-based association analyses, FUMA tested 18,818 protein-coding genes and applied a Bonferroni correction to account for multiple testing across all genes. Using 17,009 curated gene sets available in MSigDB version 2023.1.Hs (29, 30), gene-set enrichment analyses were conducted to test whether genes within predefined biological process and disease pathway gene sets showed stronger associations (i.e., lower p-values) compared to genes outside those sets.

### Heritability and genetic correlation of hypothalamic subregions with regional brain volumes

Using GWAS summary statistics from the meta-analysis, SNP-based heritability (h²_SNP) was estimated with LDSC (31) for mammillary body volume and volumes of other hypothalamic subregions using HapMap3 SNPs. Genetic correlations were subsequently calculated between the meta-analysis GWAS results for mammillary body volume and volumes for adjacent hypothalamic subregions. In addition, genetic correlation analyses were performed to compare our univariate meta-analysis GWAS summary statistics with previously published GWAS of hypothalamic subregions (5). Genetic correlation was also estimated with 101 regional brain volumetric measures (32). For this analysis, mammillary body volume GWAS summary statistics additionally adjusted for total brain volume (TBV) were used.

### Genetic correlation with traits and diseases

We estimated genetic correlations using LDSC between mammillary body volume, and hypothalamic subregions and a range of traits and diseases, including body mass index (BMI) (33), cognitive performance (34), daytime napping (35), chronotype (36), sleep duration (35), neurodegenerative diseases including Alzheimer’s disease (37) Parkinson’s disease, dementia and AvoHILMO-defined dementia (38), neurological symptoms including memory impairment (39), migraine with aura (40) and abnormal involuntary movement (40), psychiatric disorders including major depressive disorder (41) , schizophrenia (42) disorders of psychological development (38), habit and impulse disorders (38), mental and behavioral disorders due to opioids, stress-related disorder (38).

### Two-sample Mendelian Randomization

Using the TwoSampleMR R package (v0.5.7), bidirectional MR analyses were conducted to investigate the causal associations between mammillary body volume and the traits and diseases examined in the genetic correlation analyses. To minimize potential sample overlap arising from previous GWAS that included UKBB participants, GWAS for daytime napping and sleep duration were re-run in the UKBB using previously described models. For MDD, summary statistics excluding UKBB participants (pgc-mdd2025_no23andMe-noUKBB_eur_v3-49-24-11.txt) were used. For systolic and diastolic blood pressure, summary statistics from a large independent GWAS conducted in the Million Veteran Program (MVP) were used. The primary method for causal estimation was the inverse-variance weighted (IVW) (43) regression approach. Sensitivity analyses were performed using additional MR methods, including the weighted median, weighted mode (44), and MR-Egger regression (45). For the IVW and MR-Egger models, heterogeneity among SNP instruments was assessed using Cochran’s Q test (46). When significant heterogeneity was detected, outlier instruments were identified and removed using radial MR (47). To assess horizontal pleiotropy, the MR-Egger intercept test was applied. Independent genetic instruments were selected using linkage disequilibrium (LD) clumping with a 10,000 kb window and an r² threshold of 0.001, retaining SNPs reaching genome-wide significance (p < 5 × 10) based on the European ancestry reference panel from the 1000 Genomes Project. SNP instruments with an F-statistic < 10 were excluded to minimize weak instrument bias (48, 49).

### Functional validation through gene expression analyses

Candidate genes were identified through positional, expression quantitative trait loci (eQTL), and chromatin interaction mapping implemented in FUMA. The eQTL analyses were performed for the independent lead SNPs (r^2^ < 0.1) and candidate SNPs in linkage disequilibrium with the lead SNPs (r^2^ > 0.6) using residualized gene expression levels. Gene expression levels were adjusted for age, sex, the first 10 genetic principal components, red and white blood cell counts, the relative fractions of basophils, eosinophils, lymphocytes, monocytes, and neutrophils, and batch effect. Statistical significance was defined using an FDR threshold of < 0.05. To functionally validate prioritized candidate genes, association between mammillary body volume (dependent variable) with residualized gene expression levels (independent variables) were assessed using linear regression models. Gene expression residuals were generated after adjusting for the same covariates as in the eQTL analysis, namely age, sex, batch effects, red and white blood cell counts, and the proportions of basophils, eosinophils, lymphocytes, monocytes, and neutrophils. A nominal significance threshold of p < 0.05 was applied for this step. For gene–SNP combinations showing significant SNP–gene and gene–mammillary body volume associations, mediation analyses were performed using structural equation modeling in the R package *lavaan* (v0.6-11). A standard three-variable mediation framework was used as: gene expression ∼ a × SNP, mammillary body volume ∼ b × gene expression + c′ × SNP, with path *a* representing the effect of the SNP on gene expression, path *b* representing the effect of gene expression on mammillary body volume conditional on the SNP effect, and path *c*′ the direct effect of the SNP on mammillary body volume conditional on gene expression. The indirect effect was calculated as *a × b*, and the total effect as *a × b + c*′. Models were fitted using the sem() function in *lavaan*, and confidence intervals were estimated using 1,000 bootstrap resamples. For mediation analyses, gene expression residuals were adjusted for age, sex, batch effects, and blood cell counts, while mammillary body volume residuals were adjusted for age, age², sex, eTIV, genetic principal components and frontal, parietal, temporal, and occipital cortical volumes.

### Functional validation through protein expression analyses

For protein-level validation, we analyzed circulating plasma proteins, focusing on proteins linked to lead SNPs (r^2^ < 0.1) and candidate SNPs (r^2^ > 0.6) mapped to candidate genes using SEER and OLINK proteomics platforms. pQTL analyses were conducted with models adjusted for age, sex, batch effects, the first 10 genetic principal components, and the first principal component of protein expression (for SEER), as well as estimated glomerular filtration rate (eGFR). For OLINK data, adjustment for plate effects and protein-expression principal components was not required. To evaluate potential causal effects of circulating protein levels on mammillary body volume, we additionally performed Mendelian randomization using publicly available pQTL summary statistics from the deCODE Genetics large-scale plasma proteomics study as the exposure and the mammillary body volume GWAS meta-analysis as the outcome.

### Colocalization and Mendelian randomization analysis

We performed colocalization analyses to determine whether genetic variants associated with candidate genes showing significant mediation effects between SNPs and mammillary body volume shared causal variants with eQTLs identified in whole blood and brain tissues from GTEx v8. Variants located within ±500 kb of the lead SNPs were extracted and merged with GTEx v8 eQTL summary statistics. Colocalization analyses were conducted using the coloc.abf() function implemented in the R package coloc (50). This method estimates posterior probabilities for five hypotheses: H0, neither trait is associated with the region; H1 and H2, only one of the two traits is associated with this region; H3, both traits are associated but with distinct causal variants; and H4, both traits are associated and share a common causal variant. The prior probability of a shared causal variant (H4) was set to 1×10 . Evidence for colocalization was considered strong when the posterior probability for H4 (PP.H4) exceeded 0.75. To investigate potential causal relationships between gene expression and mammillary body volume, we also performed two-sample MR analyses using the R package TwoSampleMR as described above. eQTLs from whole blood and brain tissues were used as instrumental variables for gene expression (exposure), while mammillary body volume was used as the outcome. Exposure and outcome datasets were harmonized to align effect alleles and ensure consistent effect directions.

## RESULTS

### Sample characteristics

We included genetically unrelated participants from both cohorts, with sample sizes of 38,064 and 4,860 individuals from UKBB and the Rhineland Study, respectively (**Supplementary Table 1**). The average age was higher among UKBB (64.5 ± 7.7 years) than Rhineland Study participants (55.6 ± 13.4 years). The mean hypothalamic volume was slightly larger in participants of the Rhineland Study (1125.9 ± 105.5 mm^3^) compared to those of UKBB (1103.5 ± 119.7 mm^3^). In contrast, the mean mammillary body volume was slightly larger in UKBB (217.1 ± 29.8 mm^3^) as compared to the Rhineland Study (210 ± 26.9 mm^3^) participants.

### Univariate GWAS of mammillary bodies

The mammillary body volume GWAS identified 34 genomic risk loci, of which 26 were novel, after adjustment for adjacent cortical brain structures (**Table 1**, **Figure 1A, Supplementary Table 2**). These 34 loci were represented by 40 independent lead SNPs (r² < 0.1), including 28 novel lead variants (**Supplementary Table 3**). The strongest association signal among the lead SNPs was observed on chromosome 7 and mapped to the *SNORA73* locus (rs77126132, P = 5.17 × 10□²□ , β = 0.13, SE = 0.01). Other highly significant loci included variants near *NAMPTL* on chromosome 10 (rs55747874, P = 1.76 × 10□²³, β = 0.161, SE = 0.016), *RP11-443I9.1* (rs17048681, P = 3.03 × 10□²³, β = −0.074, SE = 0.007), *BICD1* on chromosome 12 (rs7298381, P = 1.15 × 10□²□, β = 0.068, SE = 0.007) and *RP11-210L7.1* (rs1078870, P = 2.43 × 10□¹□, β = −0.086, SE = 0.010) on chromosome 12, *ESRRG* on chromosome 1 (rs113229445, P = 4.68 × 10□¹□, β = −0.160, SE = 0.019), and *RFC3P1* on chromosome 3 (rs4678518, P = 6.75 × 10□¹□, β = −0.063, SE = 0.008). Several of these top loci have previously been associated with hypothalamic and posterior hypothalamic volumes (5). Among the novel lead SNPs, variants were mapped to nearby genes including *UBE2D1*, *ONECUT2*, *SIM1*, *ZFHX3*, *KIAA1462*, *CSDC2*, *XRCC4*, *DNAJC6*, and *PITPNM1* (**Table 1, Supplementary Table 4**). In the independent Rhineland Study cohort, 20 lead SNPs representing 20 genomic risk loci identified in the discovery cohort showed directional consistency and nominal replication (P < 0.05) (**Table 1**, **Figure 1B, Supplementary Table 2**). Gene-based and gene-set analysis performed using MAGMA are reported in **Supplementary Table 5** & **6**.

**Table 1:** Genomic loci associated with mammillary body volume in the discovery cohort with corresponding estimates from the replication cohort.

| rsID | Chr | Position | Other allele | Effect allele | MAF | UKB-P | UKB-BETA | UKB-SE | RS-BETA | RS-SE | RS-P | Nearest gene |
|---|---|---|---|---|---|---|---|---|---|---|---|---|
| rs7528114* | 1 | 65805144 | A | G | 0.43 | 4.93e-09 | 0.04 | 0.007 | -0.007 | 0.021 | 0.74 | *DNAJC6* |
| rs74993515* | 1 | 98336754 | T | G | 0.07 | 2.48e-08 | 0.08 | 0.014 | 0.069 | 0.04 | 0.0901 | *DPYD* |
| rs113229445 | 1 | 217073555 | G | A | 0.04 | 4.68e-17 | -0.16 | 0.019 | -0.247 | 0.055 | 8.06e-06 | *ESRRG* |
| rs73101332* | 1 | 232772373 | C | T | 0.14 | 4.59e-08 | -0.06 | 0.011 | -0.1 | 0.03 | 8.50e-04 | *RNU6-1211P* |
| rs17048681* | 2 | 57365335 | T | G | 0.35 | 3.03e-23 | -0.07 | 0.007 | -0.117 | 0.021 | 1.59e-08 | *RP11-443I9.1* |
| rs921152* | 2 | 61563275 | T | C | 0.46 | 5.44e-09 | -0.04 | 0.007 | - | - | - | *USP34* |
| rs7591656* | 2 | 64662732 | C | T | 0.29 | 4.35e-10 | 0.05 | 0.008 | 0.044 | 0.022 | 0.047 | *AC008074.3* |
| rs4678518 | 3 | 36125614 | G | A | 0.35 | 6.75e-17 | -0.06 | 0.008 | - | - | - | *RFC3P1* |
| rs624354* | 4 | 20300192 | G | A | 0.09 | 4.17e-08 | 0.06 | 0.012 | 0.156 | 0.033 | 1.67e-06 | *SLIT2* |
| rs56029134* | 4 | 112497988 | G | A | 0.14 | 3.14e-11 | 0.07 | 0.011 | 0.055 | 0.029 | 0.0637 | *RP11-255I10.1* |
| rs7661046 | 4 | 145260495 | T | G | 0.28 | 5.76e-12 | -0.06 | 0.008 | -0.059 | 0.023 | 0.0101 | *RP11-361D14.2* |
| rs2386242* | 5 | 82487199 | G | A | 0.42 | 3.60e-09 | -0.04 | 0.007 | -0.018 | 0.02 | 0.368 | *XRCC4* |
| rs1833753* | 5 | 123395938 | G | C | 0.24 | 6.35e-09 | -0.05 | 0.008 | 0.018 | 0.023 | 0.445 | *HMGB1P29* |
| rs114462827* | 6 | 26610799 | C | A | 0.01 | 1.81e-09 | 0.29 | 0.048 | 0.285 | 0.088 | 0.00130 | *RP11-457M11.6* |
| rs2535666* | 6 | 94784471 | G | A | 0.33 | 1.66e-11 | -0.05 | 0.008 | -0.031 | 0.022 | 0.152 | *RP3-463P15.1* |
| rs3798483* | 6 | 100909653 | T | C | 0.18 | 6.49e-11 | -0.06 | 0.009 | -0.034 | 0.025 | 0.179 | *SIM1* |
| rs1057947* | 7 | 8152979 | G | C | 0.05 | 2.54e-08 | -0.09 | 0.016 | -0.007 | 0.042 | 0.87 | *ICA1* |
| rs77126132 | 7 | 54966738 | G | A | 0.12 | 5.17e-26 | 0.13 | 0.013 | 0.149 | 0.036 | 3.14e-05 | *SNORA73* |
| rs4907380 | 8 | 142645621 | C | T | 0.34 | 1.81e-08 | -0.05 | 0.008 | -0.012 | 0.022 | 0.592 | *AC138647.1* |
| rs3944154* | 9 | 118340816 | T | C | 0.06 | 2.32e-08 | 0.08 | 0.014 | 0.062 | 0.042 | 0.141 | *RP11-284G10.1* |
| rs1887317* | 10 | 30407169 | T | A | 0.2 | 2.08e-10 | -0.06 | 0.009 | -0.06 | 0.027 | 0.0235 | *KIAA1462* |
| rs1326949* | 10 | 36180586 | A | G | 0.16 | 4.07e-09 | 0.06 | 0.01 | 0.086 | 0.029 | 0.00261 | *RP11-309N24.1* |
| rs55747874 | 10 | 36807887 | C | G | 0.04 | 1.76e-23 | 0.16 | 0.016 | 0.14 | 0.043 | 0.00122 | *NAMPTL* |
| rs12358567* | 10 | 60118624 | G | A | 0.21 | 1.07e-17 | 0.07 | 0.009 | 0.045 | 0.025 | 0.069 | *UBE2D1* |
| rs11604662* | 11 | 67268048 | G | A | 0.33 | 4.94e-09 | -0.04 | 0.008 | -0.011 | 0.021 | 0.605 | *PITPNM1* |
| rs7298381 | 12 | 32530257 | G | T | 0.46 | 1.15e-20 | 0.07 | 0.007 | 0.084 | 0.02 | 3.11e-05 | *BICD1* |
| rs1078870 | 12 | 102958920 | C | T | 0.16 | 2.43e-17 | -0.09 | 0.01 | -0.094 | 0.027 | 5.00e-04 | *RP11-210L7.1* |
| rs4788696* | 16 | 73070310 | A | C | 0.15 | 1.77e-10 | -0.06 | 0.01 | -0.092 | 0.028 | 8.17e-04 | *ZFHX3* |
| rs9652941* | 18 | 1986227 | C | T | 0.38 | 2.11e-14 | 0.06 | 0.007 | 0.075 | 0.021 | 3.51e-04 | *RP11-161I6.2* |
| rs7226604* | 18 | 37950651 | C | T | 0.35 | 2.79e-10 | -0.05 | 0.008 | -0.066 | 0.021 | 1.88e-03 | *RNU7-145P* |
| rs655313* | 18 | 55157989 | T | G | 0.3 | 3.54e-14 | 0.06 | 0.008 | - | - | - | *ONECUT2* |
| rs306755* | 20 | 3099752 | T | C | 0.43 | 3.58e-08 | -0.04 | 0.007 | -0.042 | 0.02 | 0.0355 | *UBOX5-AS1:UBOX5* |
| rs73136133* | 20 | 51345737 | A | G | 0.23 | 4.11e-13 | 0.06 | 0.009 | 0.068 | 0.025 | 0.00539 | *RP4-715N11.2* |
| rs6002408* | 22 | 41967870 | C | T | 0.21 | 3.16e-10 | 0.06 | 0.009 | 0.003 | 0.026 | 0.911357 | *CSDC2* |

\*) Novel genomic loci.
Abbreviations: Chr. = chromosome, MAF = minor allele frequency; UKB-P, UKB-BETA, UKB-SE and RS-P, RS-BETA and RS-SE refer to the p-value, effect size and standard error of the single nucleotide polymorphism in UKBB (discovery cohort) and the Rhineland Study (replication cohort), respectively. P = association P-value; BETA = estimated effect size; SE = standard error; SNP = single-nucleotide polymorphism; UKB = UK Biobank; RS = Rhineland Study.

**Figure 1:**
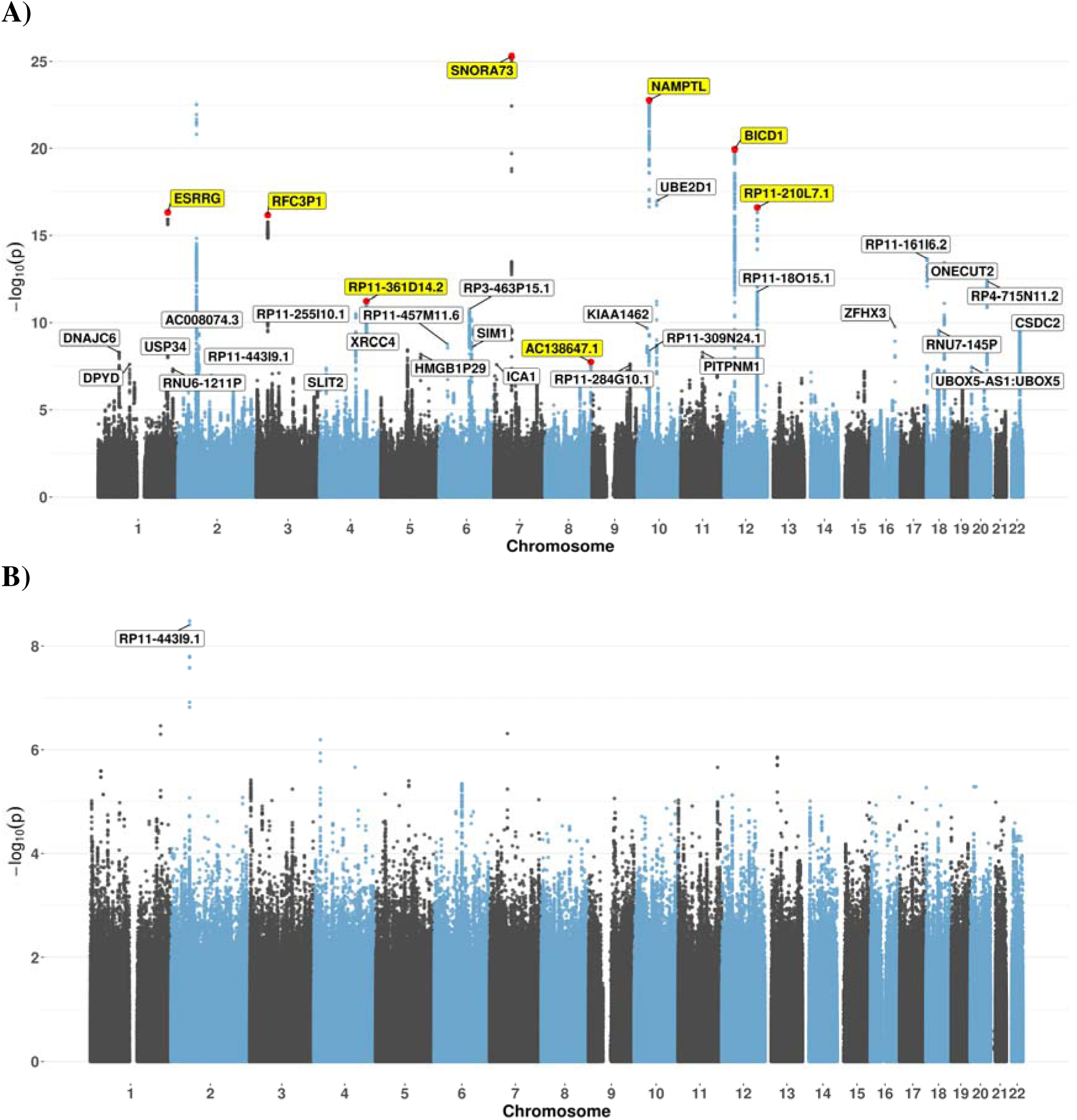
Manhattan plots showing the results in the discovery GWAS in UKBB (A), and the replication GWAS in the Rhineland Study (B). Genome-wide significant loci (p-value < 5E-8) are labeled with genes assigned through positional mapping of the lead SNPs (r² < 0.1). Genes highlighted in yellow represent loci reported in a previous GWAS.

### Multivariate GWAS of hypothalamic subregions

Multivariate GWAS of hypothalamic subregions, including the mammillary bodies, identified 55 genomic loci, of which only 15 had been reported previously (5) (**Figure 2, Supplementary Table 7**), represented by 64 independent lead SNPs (**Supplementary Table 8**). One of the replicated loci, *ADAMTS8* (mapped to SNP rs3740888), represented a top locus in both the current and previous multivariate GWAS (5). Among the novel loci were *AC005008.2*, *RP3-495K2.3*, *STRN*, *UBE2D1*, *COL11A1*, *WNT3*, *C16orf95*, *VSTM4*, *ASPG*, *C2orf47*, *RP11-161I6.2*, *ROBO1*, *GPATCH1*, *ONECUT2*, *AFAP1*, *EPHA4*, *KIAA1462*, *LINC00992*, *RSPO2*, and *RNU7-145P* (**Supplementary Table 9**).

**Figure 2:**
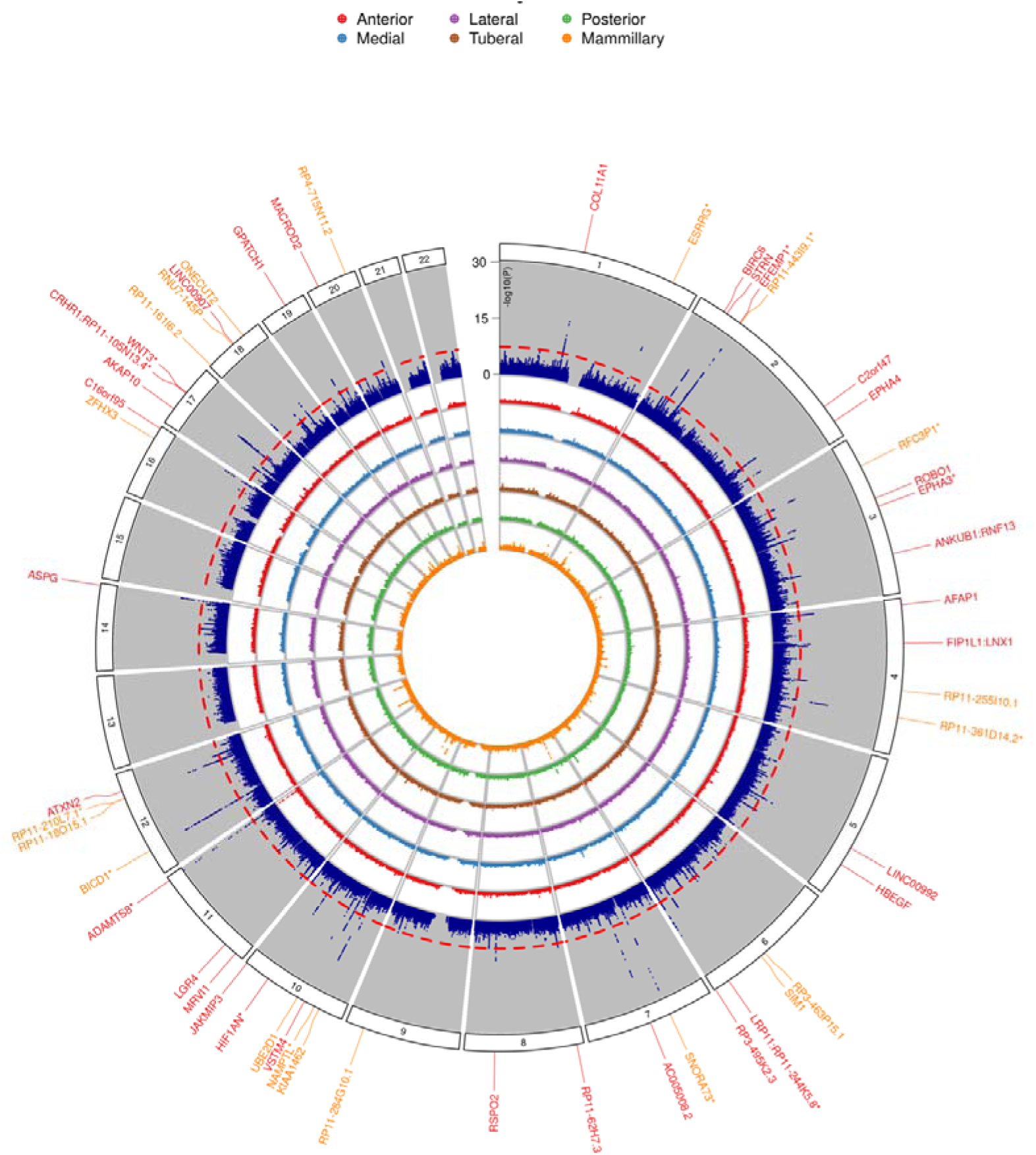
Circular Manhattan plot of the multivariate GWAS of all hypothalamic subregions. The outer blue circle represents the results of the multivariate GWAS analysis. Significant genomic loci are annotated with genes mapped to the corresponding lead SNPs. An asterisk (*) indicates loci that were also reported in a previous GWAS of hypothalamic subregions^6^. Genes highlighted in orange represent loci that also reached genome-wide significance in the univariate mammillary body GWAS.

Of the 40 novel loci, 31 reached genome-wide significance in the univariate GWAS of individual hypothalamic subregions. Nineteen showed their strongest associations with the mammillary bodies, while four were primarily associated with the anterior hypothalamus, three with the medial hypothalamus, one with the tuberal hypothalamus, and four with the posterior hypothalamus. Among loci shared across univariate GWAS, *SNORA73* was associated with both the mammillary bodies and the posterior hypothalamus (**Supplementary Table 7**). MAGMA gene-based analysis identified 117 genes, with *AC007382*.1 (6.59 × 10^1^) and *STH* (6.87 × 10^1^) showing the strongest associations (**Supplementary Table 10**). Gene-set analysis of 17,009 pathways identified nine pathways that survived Bonferroni correction, with GOBP_CARTILAGE_DEVELOPMENT showing the strongest association (P.bon < 0.0001) (**Supplementary Table 11**). Using positional and functional gene mapping strategies, we identified 28 genes consistently implicated across all four gene mapping strategies (**Supplementary Figure 1A, B**). Using GTEx v8 tissue enrichment analysis across 53 tissue types revealed that these genes are differentially expressed in the anterior cingulate cortex, frontal cortex and hypothalamus (**Supplementary Figure 1C**).

### Meta-analysis GWAS of mammillary body volume

The meta-analysis GWAS combining summary statistics from UKBB and the Rhineland Study for mammillary body volume replicated 29 genome-wide significant genomic risk loci identified in the discovery GWAS and identified 15 additional genomic loci (**Figure 3, Table 2, Supplementary Table 12**). These loci included *CAPZA1*, *EXOC4*, *STMN4*, *SFRP1*, *RSPO2*, *ZNF462*, *KIAA1328*, *UBOX5:FASTKD5* and *NHP2L1*. In total, among 44 genomic loci identified, 35 were novel. Significant heterogeneity was observed only for rs6789476 mapped to *AC026188*.*1* (I² = 84.4%, P= 0.011), while rs3790598 mapped to *CAPZA1,* showed substantial but borderline heterogeneity (I² = 73.1%, P = 0.054). Across these 44 genomic loci, 53 independent lead SNPs (r² < 0.1) were identified (**Supplementary Table 13**), which were in linkage disequilibrium (r² > 0.6) with 5,921 candidate SNPs (**Supplementary Table 14**).

**Figure 3:**
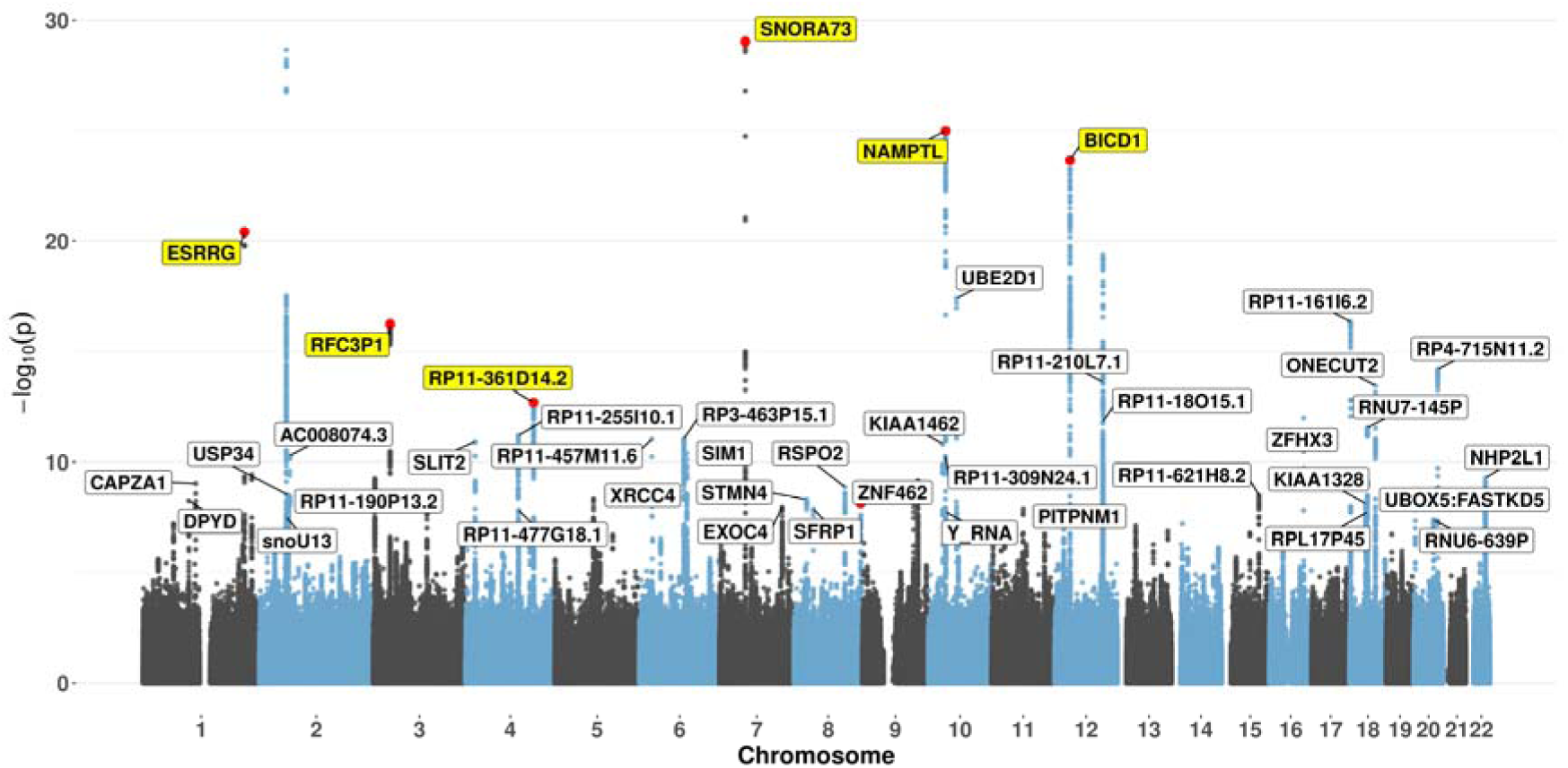
Manhattan plots showing the results of the meta-GWAS of mammillary body volume in UKBB and the Rhineland Study combined. Genome-wide significant loci (p-value < 5E-8) are labeled with genes assigned through positional mapping of lead SNPs (r² < 0.1). Genes highlighted in yellow represent loci that were also reported in a previous GWAS.

**Table 2:**
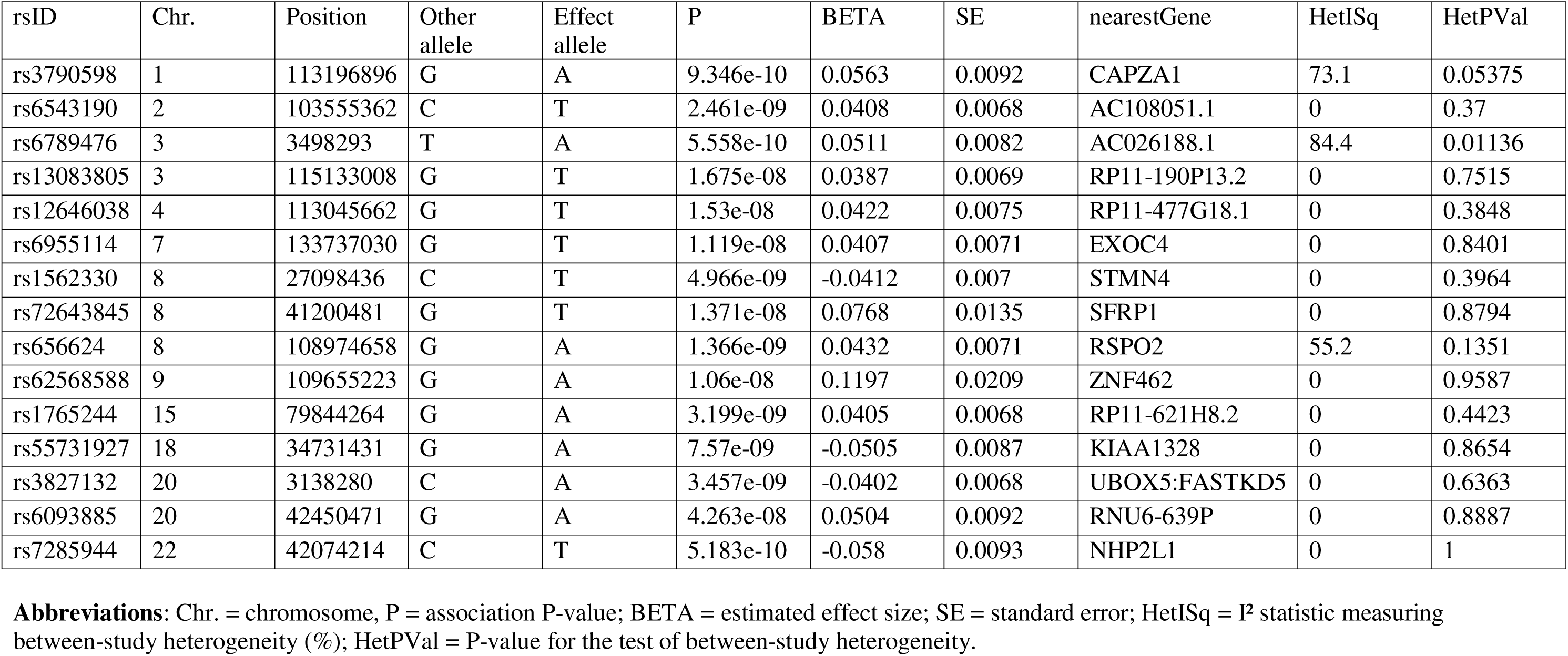
Newly identified genomic loci associated with mammillary body volume in the meta-GWAS analysis.

### Gene and functional mapping of mammillary body volume meta-GWAS

Using independent lead SNPs and candidate SNPs in LD (r² > 0.6) with the lead variants based on meta-analysis GWAS summary statistics, gene mapping was performed through multiple strategies (**Supplementary Table 14**). Positional mapping identified 47 genes, eQTL mapping identified 57 genes, and chromatin interaction mapping identified 178 genes. In addition, MAGMA gene-based analysis identified 47 genes that reached gene-wide significance (P < 2.66 × 10) (**Supplementary Table 15**). By overlapping genes identified across all mapping strategies, we prioritized 13 genes, namely *C2orf74*, *CCDC134*, *CCDC53*, *DESI1*, *KIAA1841*, *MEI1*, *MOV10*, *NHP2L1*, *PARPBP*, *RSPO2*, *TOB2*, *XPO1* and *XRCC4* that may have functional relevance to mammillary body volume (**Figure 4**). Positional gene-set enrichment analysis further identified significant enrichment of two cytogenetic regions, chr22q13 and chr2p15 (**Figure 4C**). The chr22q13 region included *TOB2*, *DESI1*, *NHP2L1*, *MEI1* and *CCDC134*, whereas chr2p15 region included *KIAA1841* and *XPO1*.

**Figure 4:**
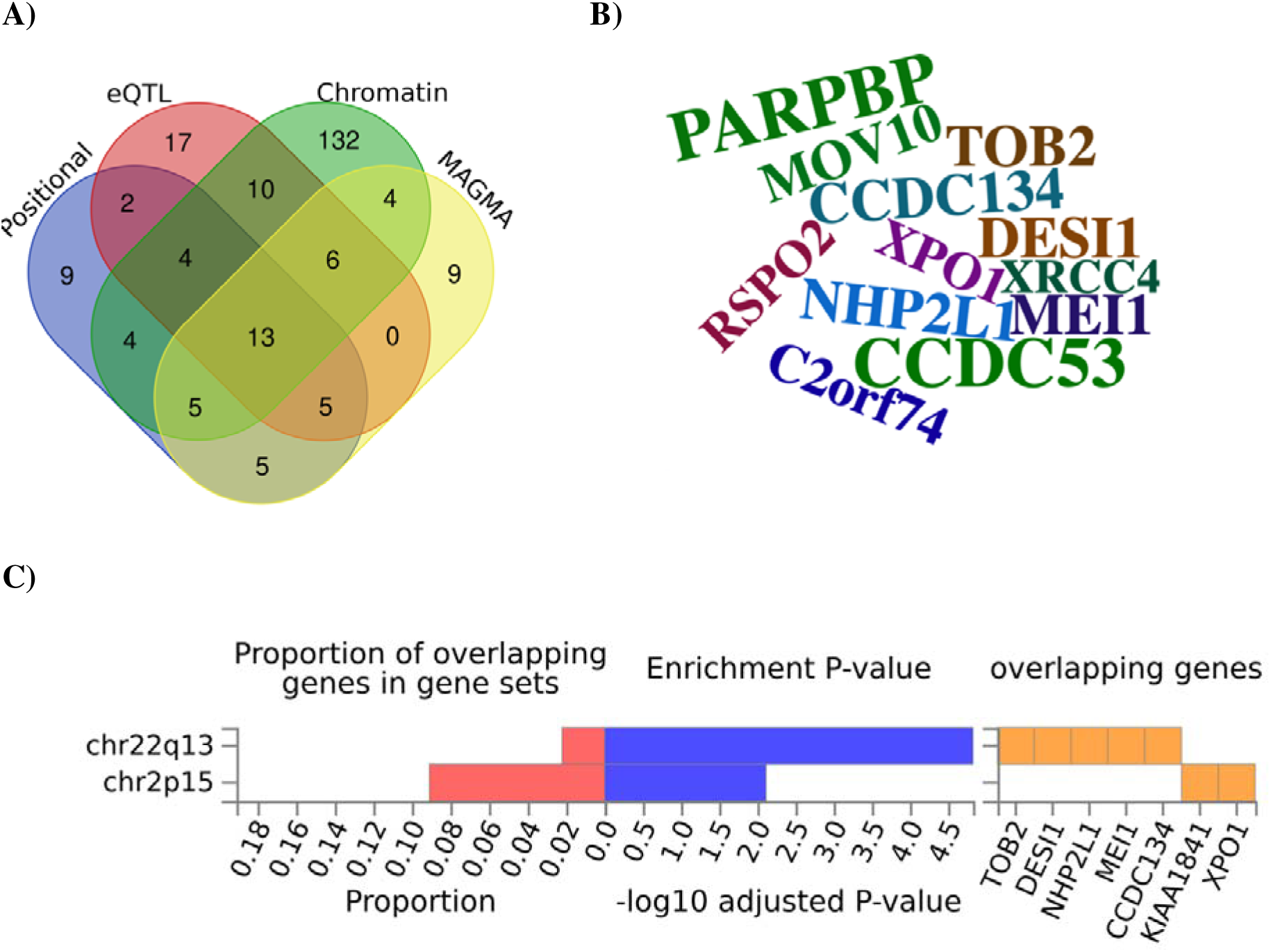
Gene mapping. **A)** Gene mapping of the meta-GWAS results of mammillary body volume based on positional and functional annotation of lead (r^2^<0.1) and candidate SNPs (r^2^>0.6) via positional, expression quantitative trait loci (eQTL), chromatin interaction and MAGMA gene-based mapping. **B)** Word cloud displaying genes implicated by all four strategies, with word size proportional to the -log (p) values from the GWAS. **C)** Positional gene-set enrichment analysis of candidate genes showing significantly enriched cytogenetic regions. The left panel displays the proportion of overlapping genes within each enriched gene set, the middle panel shows the enrichment significance (−log adjusted P-value), and the right panel lists the overlapping genes contributing to each enriched gene set.

### SNP heritability and genetic correlation of mammillary body volume meta-GWAS with other hypothalamic subregions

Using genome-wide GWAS summary statistics from the meta-analysis, LD Score Regression (LDSC) estimated the SNP-based heritability (h²) of mammillary body volume to be 0.29 (SE = 0.021) (**Supplementary Table 17, Figure 5A**). Genetic correlation analyses demonstrated significant positive correlations between mammillary body volume and the whole hypothalamus (rg = 0.6354, SE = 0.0259, P = 4.08 × 10□¹³³, FDR-adjusted P = 2.45 × 10^-132^), posterior hypothalamus (rg = 0.4125, SE = 0.0383, P = 5.31 × 10□²□, FDR-adjusted P = 1,59 × 10^-26^), lateral hypothalamus (rg = 0.3904, SE = 0.0515, P = 3.39 × 10□¹□, FDR-adjusted P = 6.78 × 10^-14^), and medial hypothalamus (rg = 0.1621, SE = 0.0474, P = 6.00 × 10 , FDR-adjusted P = 0.0009). A modest negative genetic correlation was observed with the tuberal hypothalamus (rg = −0.1277, SE = 0.0557, P = 2.18 × 10□², FDR-adjusted P = 0.026) (**Supplementary Table 18, Figure 5A**). Genetic correlations were also estimated between mammillary body volume and hypothalamic subregions reported in previous GWAS. Significant positive correlations were observed with anterior superior (rg = 0.3641, SE = 0.0567, P = 1.34 × 10□¹□, FDR-adjusted P = 1 × 10^9^), anterior inferior (rg = 0.162, SE = 0.0566, P = 0.0042, FDR-adjusted P = 0.021), posterior (rg = 1.0080, SE = 0.0215, P < 1 × 10=□³□□, FDR-adjusted P < 1 × 10□³□), tuberal inferior (rg = 0.3564, SE = 0.0538, P = 3.50 × 10□¹, FDR-adjusted P = 3.5 × 10^10^), tuberal superior (rg = 0.3577, SE = 0.0564, P = 2.23 × 10□¹□, FDR-adjusted P = 1.34 × 10□^9^), and whole hypothalamus (rg = 0.7953, SE = 0.0318, P = 1.24 × 10□¹³□, FDR-adjusted P = 1.86 × 10□^136^) (**Supplementary Table 20, Figure 5B**).

**Figure 5:**
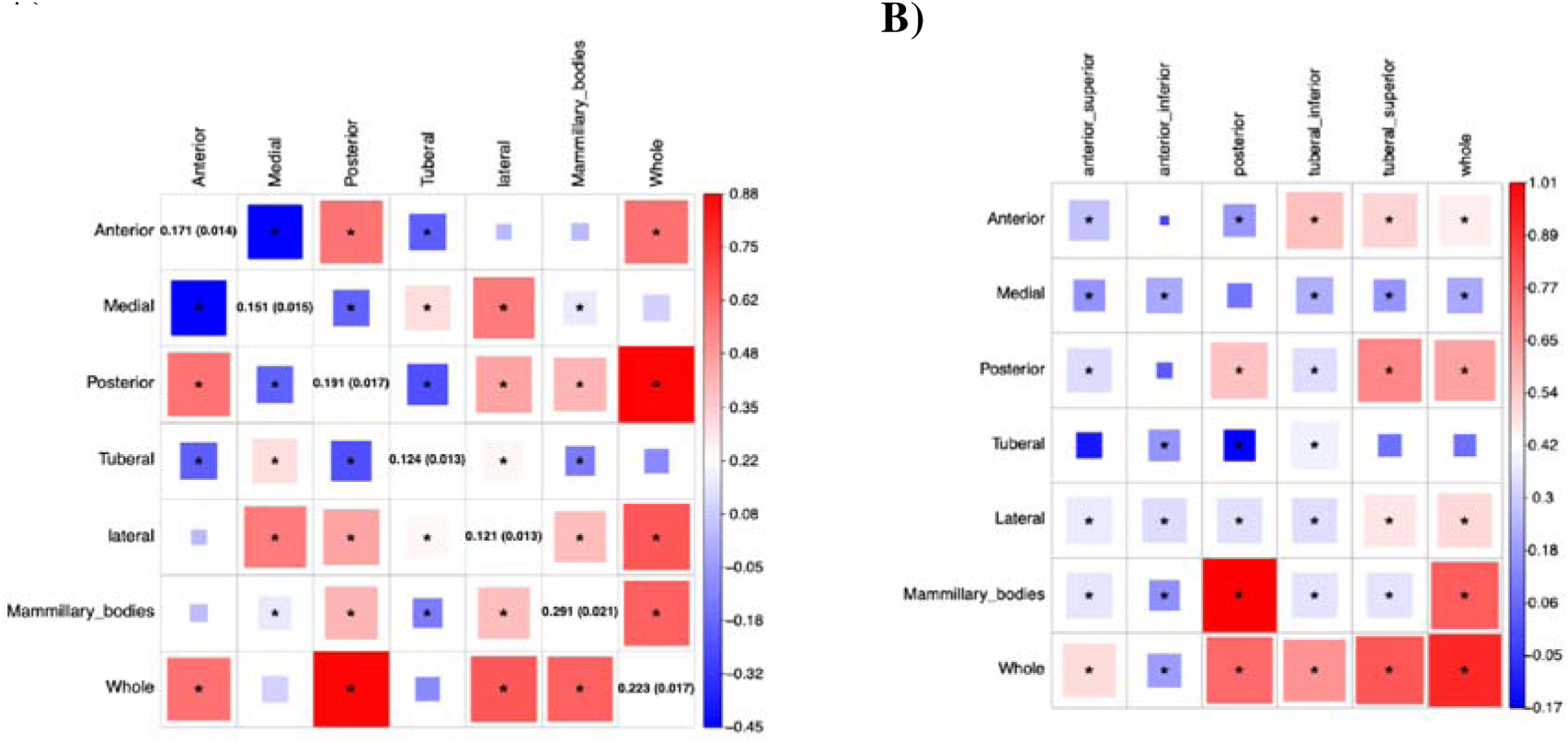
Linkage disequilibrium score regression analyses. **A)** SNP-based heritability (h²_SNP_) and genetic correlations between mammillary body volume and other hypothalamic subregions in the GWAS meta-analysis. **B)** Genetic correlations between mammillary body volume and other hypothalamic subregions based on summary statistics from a previously published GWAS of hypothalamic subregional volumes. *Indicates FDR significant.

### Genetic correlation of mammillary body volume meta-GWAS with 101 other brain structures

We estimated genetic correlations between mammillary body volume and 101 regional brain volumetric measures. Mammillary body volume showed significant positive genetic correlations with multiple regional brain volumes, including the right ventral diencephalon (rg = 0.3333, SE = 0.0406, FDR-adjusted P = 2.35 × 10^14^) and left ventral diencephalon (rg = 0.3464, SE = 0.0437, FDR-adjusted P = 1.17 × 10^13^), right hippocampus (rg = 0.1938, SE = 0.0452, FDR-adjusted P = 6.09 × 10^4^), brainstem (rg = 0.1928, SE = 0.0458, FDR-adjusted P = 6.39 × 10^4^), left thalamus proper (rg = 0.2207, SE = 0.0571, FDR-adjusted P = 1.12 × 10^3^), left hippocampus (rg = 0.1651, SE = 0.0434, FDR-adjusted P = 1.12 × 10^3^), right thalamus proper (rg = 0.2141, SE = 0.0565, FDR-adjusted P = 1.12 × 10^3^), right pallidum (rg = 0.1536, SE = 0.0402, FDR-adjusted P = 1.12 × 10^3^), left pallidum (rg = 0.1158, SE = 0.0398, FDR-adjusted P = 3.31 × 10^2^) and right cerebellar white matter (rg = 0.1323, SE = 0.0474, FDR-adjusted P = 4.38 × 10^2^). Significant negative genetic correlations were observed with total gray matter volume (rg = −0.1782, SE = 0.0466, FDR-adjusted P = 1.12 × 10^3^) and the left lateral occipital cortex (rg = −0.2289, SE = 0.0730, FDR-adjusted P = 1.72 × 10^2^) (**Supplementary Table 19, Figure 6**).

**Figure 6:**
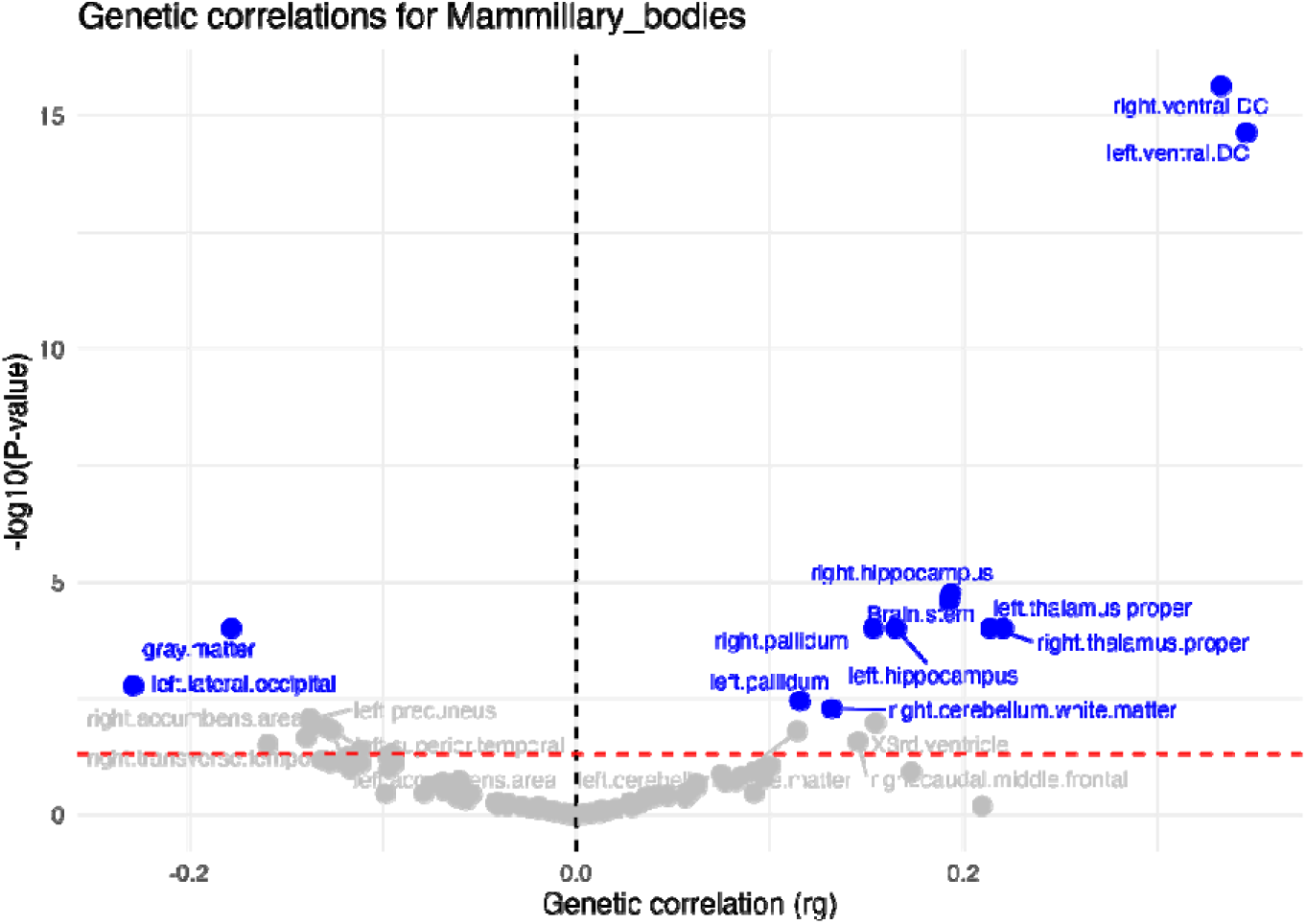
Linkage disequilibrium score regression and genetic correlation analyses of mammillary body volume and volumes of 101 other brain regions. Blue dots correspond to significant genetic correlations after correction for multiple testing (FDR).

### Genetic correlation of mammillary body volume meta-GWAS with traits and diseases

Given the important role of the hypothalamus in the regulation of a diverse array of homeostatic functions, we estimated the genetic correlation between mammillary body volume and anthropometric measures (including BMI), cognition, neurological symptoms such as memory impairment, abnormal involuntary movements and migraine with preceding neurological symptoms, neurodegenerative diseases such as Parkinson’s disease and Alzheimer’s disease, mental disorders such as all-cause dementia, MDD, disorders of psychological development, habit impulse disorders, opioid-related and severe stress related disorders, and sleep related traits such as daytime napping, chronotype and sleep duration. Among these traits, significant genetic correlations were observed for all-cause dementia (rg = -0.1579, SE = 0.0659, P = 0.0166, FDR-adjusted P = 0.0383), MDD (rg = -0.0617, SE = 0.0240, P = 0.0101, FDR-adjusted P = 0.0336), opioid related disorder (rg = -0.1767, SE = 0.0725, P = 0.0148, FDR-adjusted P = 0.0383), severe stress related disorder (rg = -0.1414, SE = 0.054, P = 0.0088, FDR-adjusted P = 0.033), disorders of psychological development (rg = 0.2509, SE = 0.1026, P = 0.0145, FRD-P = 0.0383) and abnormal involuntary movements (rg = -0.1037, SE = 0.0431, P = 0.0162, FDR-adjusted P = 0.0383) (**Supplementary Table 21, Figure 7**).

**Figure 7:**
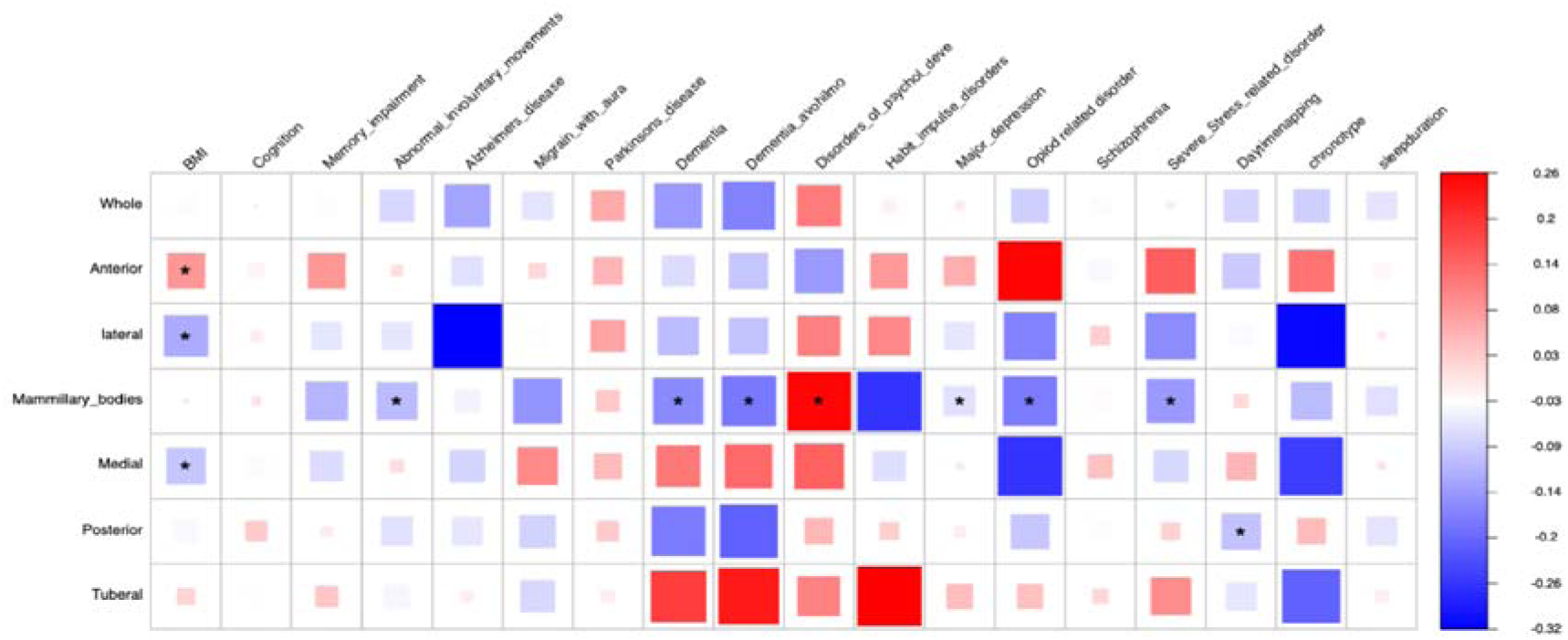
Genetic correlations between mammillary body volume and health-related traits, including neurological, neurodegenerative, psychiatric, mood, cognitive, and behavioral phenotypes. *Indicates FDR significant.

### Two-sample Mendelian Randomization

We investigated potential causal relationships between mammillary body volume and disorders that survived FDR correction in the genetic correlation analysis using bidirectional two-sample MR. Among these, only MDD showed evidence of causal association. In the forward MR analysis, the primary inverse-variance weighted (IVW) analysis showed that smaller mammillary body volume was associated with an increased risk of MDD, although the association did not reach statistical significance (IVW: β = −0.026, SE = 0.019, P = 0.176). Significant heterogeneity was detected among the genetic instruments (Cochran’s Q = 107.35, P = 1.56 × 10). After excluding outlier SNPs identified by radial MR, the association became statistically significant (IVW: β = −0.037, SE = 0.012, P = 1.72 × 10 ³, FDR-adjusted P = 0.012), indicating that a decrease in mammillary body volume was associated with an increased risk of MDD. The weighted median analysis showed a consistent direction of effect and remained statistically significant following outlier removal (β = −0.034, SE = 0.016, P = 0.035) **Figure 8A**). In the reverse MR analysis, a higher genetic liability to MDD was also associated with smaller mammillary body volume following removal of outlier SNPs (IVW: β = −0.10, SE = 0.03, P = 1.75 × 10 ³, FDR-adjusted P = 0.01). Together, these findings suggest a bidirectional association between mammillary body volume and MDD (**Supplementary Table 22, Figure 8B**).

**Figure 8:**
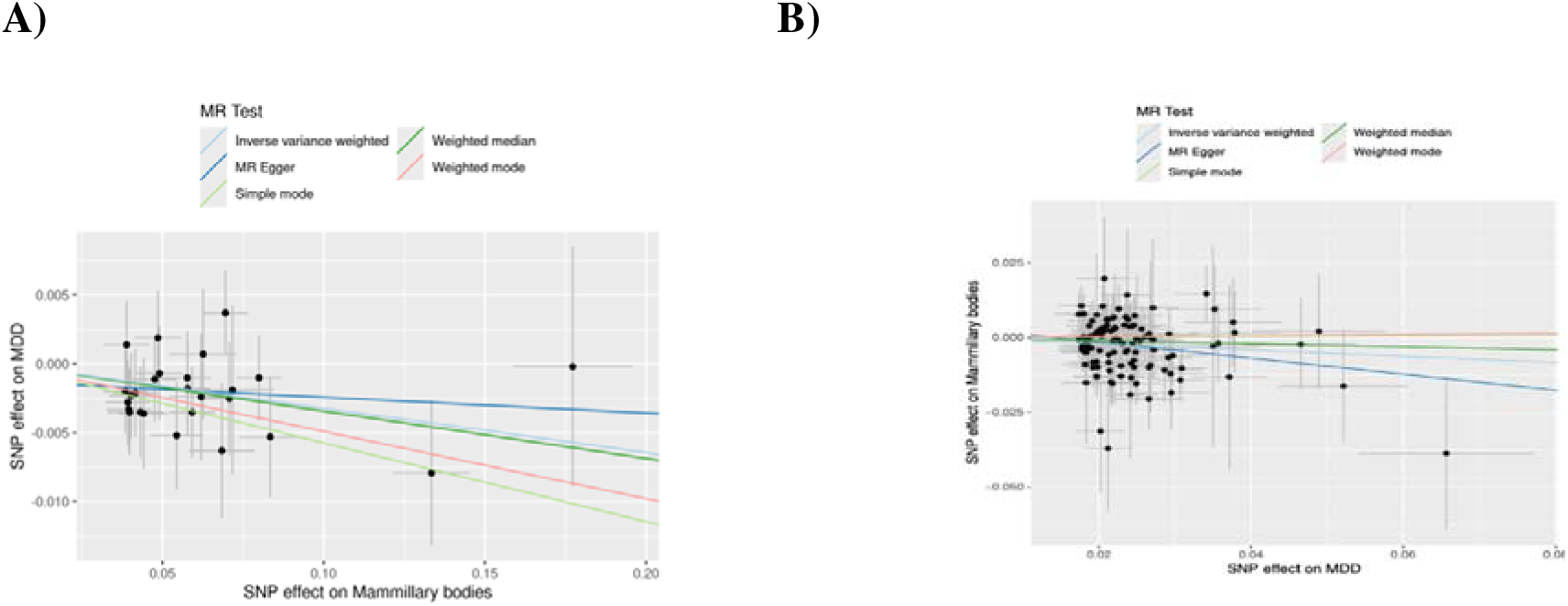
Two-sample Mendelian randomization analysis of the relationship between mammillary body volume and major depressive disorder (MDD). **A)** Forward MR analysis assessing the causal effect of mammillary body volume on MDD risk. Reverse MR analysis assessing the causal effect of MDD liability on mammillary body volume.

### Transcriptomic analysis

To further investigate the genes mapped to the lead SNPs from the meta-GWAS, we performed functional validation using individual-level transcriptomic data available in the Rhineland Study. Specifically, we leveraged 177 candidate genes originally identified through positional and functional annotation of 5921 SNPs in LD with 115 independent significant SNPs from the meta-GWAS summary statistics. Out of these, expression data on 51 genes and 1627 candidate SNPs were available in the Rhineland Study for the transcriptomic analysis. In the eQTL analysis, we identified 706 SNPs showing significant associations with the expression of 20 genes. The strongest association was observed for rs139545, which was associated with decreased expression of the *MEI1* gene (FDR-adjusted P = 1.74 × 10^55,^ P = 1.91 × 10 ^57^, β = -0.57, SE = 0.03). This was followed by rs11006122, which was associated with *UBE2D1* gene expression (FDR-adjusted P =2.41 × 10 ^52^, P = 9.46 × 10 ^54^, β = -0.36, SE = 0.02) (**Supplementary Table 23, Supplementary Figure 2A**). For genes that survived FDR correction, we further assessed the association between gene expression and mammillary body volume. Among these, *MEI1* showed a nominally significant association, with higher *MEI1* expression associated with smaller mammillary body volume (FDR = 0.42, P = 0.022, β = −0.06, SE = 0.03), followed by *GSTP1* (FDR= 0.42, P = 0.042, β = −0.061, SE = 0.03) (**Supplementary Table 24, Supplementary Figure 2B**). In addition, we performed mediation analyses using the expression level of the *MEI1* gene that was significantly associated with both mammillary body volume and its corresponding SNP (**Supplementary Table 25**). Mediation analysis revealed that the effect of 104 SNPs on mammillary body volume were significantly mediated through *MEI1* expression (indirect p < 0.05, FDR < 0.02), with the absolute ratio of the indirect effect of the total effect size ranging from 78% to over 100%.

### Causal association of *MEI1* gene with mammillary body volume

MR analysis provided support for a potential causal relationship between *MEI1* expression and mammillary body volume across multiple GTEx tissues. In whole blood, a significant negative causal association was observed (IVW β = -0.0851, SE = 0.0150, FDR-adjusted P = 2.75 × 10□□). In other brain tissues, significant causal associations were also observed, including cortex (β = -0.1111, SE = 0.0191, FDR-adjusted P = 1.70 × 10), caudate basal ganglia (β = -0.1526, SE = 0.0245, FDR-adjusted P = 3.58 × 10□□), hippocampus (β = -0.1048, SE = 0.0189, FDR-adjusted P = 4.83 × 10□□), putamen (β = -0.1422, SE = 0.0244, FDR-adjusted P = 1.70 × 10□□), hypothalamus (β = -0.0951, SE = 0.0210, FDR-adjusted P = 6.92 × 10□v), and anterior cingulate cortex BA24 (β = -0.0639, SE = 0.0117, FDR-adjusted P = 6.32 × 10□□) (**Supplementary Table 27**); these estimates were based on the Wald ratio test due to the availability of only a single SNP instrument. To further evaluate whether the same underlying genetic variants influenced both *MEI1* expression and mammillary body volume, we performed colocalization analyses across tissues. Strong evidence of colocalization (posterior probability for a shared causal variant, PP.H4 > 0.75) was observed between the mammillary body volume GWAS signal and *MEI1* eQTLs in the caudate (PP.H4 = 0.901), putamen (PP.H4 = 0.789), hypothalamus (PP.H4 = 0.783), whole blood (PP.H4 = 0.767), cortex (PP.H4 = 0.755), and hippocampus (PP.H4 = 0.755) (**Supplementary Table 26, Supplementary Figure 3**).

### Proteomic analysis

We conducted pQTL analyses for candidate SNPs and proteins encoded by 177 candidate genes using data from the SEER and OLINK proteomic platforms. In the SEER dataset, 31 proteins were available for both pQTL analyses and protein association analyses with mammillary body volume, whereas approximately 10 proteins were available in the OLINK dataset. In the SEER platform, after correction for multiple testing we identified 63 SNP-protein associations involving four proteins (GSTP1, CAPZA1, RPS6KB2 and ZC3H7B) (**Supplementary Table 28, Supplementary Figure 4A**). In the OLINK dataset, 27 SNP-protein associations involving four proteins (TNFRSF13C, SLIT2, PMCH & GSTP1) survived FDR correction (**Supplementary Table 31**). We further examined the association between circulating protein levels and mammillary body volume in both datasets. Only CAPZA1 protein levels derived from the SEER platform showed a significant association with mammillary body volume after multiple testing correction (FDR-adjusted P = 0.015, P = 0.003, β = 0.48, SE = 0.16) (**Supplementary Table 29, Supplementary Figure 4B**). Finally, we performed mediation analyses to assess whether CAPZA1 protein levels mediated the association between CAPZA1 pQTL variants and mammillary body volume. Mediation analysis revealed that seven CAPZA1 pQTL variants showed indirect effects through CAPZA1 protein levels, with a mediation effect accounting for 60.2% of the total genetic effect on mammillary body volume (**Supplementary Table 30**). To further investigate causality, we performed Mendelian randomization using the lead CAPZA1 pQTL variant as an instrumental variable. The Wald ratio analysis showed that genetically predicted higher CAPZA1 protein levels were associated with larger mammillary body volume (β = 0.35, SE = 0.15, P = 0.020).

## DISCUSSION

Leveraging data from two large-scale population-based cohorts (N=42,924), we performed the first GWAS of mammillary body volume. Our meta-GWAS identified 44 genomic loci associated with mammillary body volume, of which 35 are novel. Moreover, we found that compared to other hypothalamic subregions, mammillary body volume had the highest SNP-heritability (i.e., 29%). These findings provide a wealth of new insights into the molecular mechanisms underlying interindividual differences in mammillary body volume.

Interestingly, *SNORA73* (tagged by rs77126132) was identified as the most significant locus associated with mammillary body volume, followed by loci near *BICD1, ESRRG*, *RFC3P1*, *RP11-361D14.2*, *AC138647.1*, *NAMPTL* and *RP11-210L7.1.* Indeed, in our study, genetic correlation analyses showed significant genetic overlap between mammillary body volume and those of the posterior hypothalamus and other hypothalamic structures, consistent with previous reports (6). *SNORA73* has previously been implicated in cellular metabolism through the regulation of oxidative phosphorylation and mTOR signaling pathway (51). The identification of *SNORA73* as the most significant locus for mammillary body volume suggests that RNA-mediated metabolic regulation may contribute to structural variation in mammillary bodies. Given the role of the hypothalamus in energy homeostasis and metabolic processes (52), this finding provides a potential molecular link between metabolic regulation and the size of mammillary bodies. Notably, the mammillary bodies are highly susceptible to neuropathological damage caused by Wernicke-Korsakoff syndrome. In this syndrome, a deficiency of thiamine leads to impaired energy metabolism and causes specific neuronal damage (53). Although a direct association between *SNORA73* and thiamine metabolism has not yet been established, both are crucial in managing cellular energy pathways. Therefore, the variation in *SNORA73*-mediated regulation of cellular energy metabolism may influence the metabolic stability of the mammillary bodies, making them more susceptible to this neurological disorder. Although this hypothesis is speculative, it provides a plausible molecular link between *SNORA73*-energy regulation and mammillary bodies’ physiology. Future experimental studies are necessary to investigate this potential link. Similarly, *BICD1* (Bicaudal Cargo Adaptor 1), another associated locus, encodes an adaptor protein that regulates microtubule-based intracellular cargo transport via the dynein motor complex and is involved in cytoskeletal organization (54), which is critical for neuronal structural integrity and brain development (55). Our transcriptomic analysis revealed that the lead SNP rs7298381 and its neighboring SNPs in LD (r² > 0.6) act as eQTLs for *BICD1* in whole blood. Moreover, querying of the GTEx database further supported the role of these variants as eQTLs of *BICD1* in several other brain regions, including the caudate, hippocampus, and cerebellar hemisphere. Together, these findings support a potential role for *BICD1* in the structural development and maintenance of the mammillary bodies.

Among the genes prioritized by positional, eQTL, and chromatin interaction mapping, several have been previously implicated in neuronal development and brain structure, making them particularly plausible candidates for influencing mammillary body volume. These genes included *MOV10, RSPO2, CCDC53, XRCC4*, *TOB2, DESI1, NHP2L1, MEI1, CCDC134, KIAA1841* and *XPO1*. *MOV10* has been shown to be involved in neuronal development through cytoskeletal mRNA regulation, supporting normal brain development and function, with reduced *MOV10* expression impairing dendritic arborization in hippocampal neurons (56). *RSPO2,* which encodes R-spondin 2 protein enhances Wnt signaling through leucine-rich repeat-containing G protein-coupled receptors (LGR4-6), and is expressed in the brain (57). Moreover, *RSPO2* has been previously associated with subcortical volume and brain morphology (58), while loss of *RSPO2* in hippocampal CA3 neurons impairs synaptic plasticity and memory formation in mice (59). However, it remains to be shown how *MOV10, RSPO2* as well as the other genes associated with mammillary body volume, precisely affect the development and morphology of this structure.

Positional gene-set analysis further identified significant enrichment at chromosome 22q13 locus, which encompasses *TOB2, DESI1, NHP2L1, MEI1,* and *CCDC134*. Deletions involving the 22q13 region have been associated with Phelan-McDermid syndrome, a neurodevelopmental disorder characterized by intellectual disability, severely delayed language development, and autism spectrum features (60), further highlighting the neurodevelopmental relevance of this genomic region and its candidate genes as potential determinants of mammillary body structure.

Using individual-level transcriptomic data from the Rhineland Study, we identified 706 SNP-gene expression associations involving 20 genes. Among them, *MEI1* emerged as a particularly interesting candidate. We found genetically regulated *MEI1* expression to be associated with mammillary body volume, with additional mediation analysis revealing that *MEI1* expression partially mediates the relationship between variants in this gene and mammillary body volume. Similarly, colocalization analysis provided strong evidence for a shared causal variant influencing both mammillary body volume and *MEI1* expression across multiple tissues, including the hypothalamus, caudate, putamen, hippocampus, and whole blood. These findings suggests that *MEI1* expression in these tissues is regulated by the same genetic variants that influence mammillary body volume. *MEI1* (Meiotic Double-Strand Break Formation Protein 1) is primarily known for its role in meiotic recombination based on studies in mice (61). Our findings thus indicate that *MEI1* may represent a novel candidate gene influencing mammillary body volume and suggest that it may have previously unrecognized functions in the adult human brain, warranting further investigation.

Our proteomic analyses identified significant pQTL associations involving seven proteins (including GSTP1, CAPZA1, RPS6KB2, ZC3H7B, TNFRSF13C, SLIT2 & PMCH). Among these, CAPZA1 emerged as the most interesting protein, as circulating CAPZA1 protein levels were also significantly associated with mammillary body volume. Several CAPZA1 pQTL variants were additionally associated with CAPZA1 protein abundance and showed significant mediation effects, indicating that CAPZA1 protein levels partially mediate the relationship between variants in this gene and mammillary body volume. *CAPZA1* encodes for the alpha subunit of the F-actin capping protein, involved in maintaining cytoskeletal organization through regulation of cortical actin filaments (62), a mechanism critical for neuronal structural integrity and synaptic maintenance. Moreover, findings from mice studies suggest that CAPZA1 interacts with PCDH19, a calcium-dependent cell-adhesion protein, to affect brain development, neuron migration and differentiation (63).

Our multivariate GWAS, including the mammillary bodies alongside all other hypothalamic subregions, identified 55 genomic loci, of which 40 are novel compared to previous multivariate hypothalamic GWAS findings (5). Moreover, using gene mapping strategies, we identified 28 genes that showed significant enrichment in the hypothalamus, anterior cingulate cortex and frontal cortex, brain regions that play a critical role in neuroendocrine regulation, cognition and emotional processing.

Genetic correlation analyses revealed that mammillary body volume shares its genetic architecture with several neighboring brain structures, most notably the ventral diencephalon, hippocampus, thalamus, and pallidum. In particular, the strongest genetic correlation was observed with the ventral diencephalon bilaterally, consistent with its anatomical proximity and shared developmental origin. The genetic overlap with the hippocampus and thalamus is particularly meaningful, as these structures together with the mammillary bodies form key components of the Papez circuit, a network central to episodic memory, spatial navigation, and emotional processing. Intriguingly, we found that larger mammillary bodies showed significant genetic overlap with lower risks of dementia, MDD, opioid-related disorders and severe stress related disorder.

Interestingly, among these disorders showing significant genetic overlap with mammillary body volume, seven genes located within the mammillary body volume risk loci (i.e., *ASCC3*, *CELF4*, *ESRRG*, *EXOC4*, *SIM1*, *Y_RNA* and *ZFHX3*) have previously been implicated in the pathogenesis of depression, suggesting the presence of shared biological pathways between mammillary body structure and MDD. Indeed, through two-sample MR we found evidence for a bidirectional relationship between mammillary body volume and MDD risk. On the one hand, genetic instruments associated with mammillary body volume were associated with an increased risk of depression, suggesting that reduced mammillary body volume may contribute to depression susceptibility. On the other hand, higher genetic liability to MDD was also associated with reduced mammillary body volume, suggesting depression-related biological processes may also contribute to structural changes in mammillary bodies. Previous large-scale imaging studies have already shown that the hippocampus is smaller in people with depression (64). While most research has focused on the hippocampus, our study shows that the mammillary bodies, which connect directly to the hippocampus, also share a causal link with depression. Although these two regions are closely connected, our findings highlight the mammillary bodies as a distinct genetic factor in depression, paving the way for future studies to look at how these structures interact as a network.

Our study has several limitations. First, the sample size of our discovery GWAS, while substantial, was modest compared to GWAS of other brain structures. Nevertheless, we identified novel genetic loci associated with mammillary body volume, a large proportion of which were also replicated in the independent Rhineland Study cohort, providing novel genetic insights into mammillary body structure through meta-analysis. Second, we were unable to assess whether the identified loci are associated with temporal changes in mammillary body volume, which would be valuable for identifying genetic determinants of age-related volumetric changes. Incorporating longitudinal data on mammillary body volume in future GWAS studies would overcome this limitation and facilitate the identification of loci associated with changes over time. Finally, our GWAS was restricted to individuals of European ancestry, which limits the generalizability of our findings to other populations. Future large-scale multi-ancestry GWASs are warranted to further help assess the heterogeneity of genetic effects and identify additional genetic determinants of mammillary body size.

In this study, we identified numerous genetic loci associated with volumes of the mammillary bodies, which showed a highly heritable and distinct genetic architecture within the hypothalamus. Through integrative multi-omics analyses, we pinpointed several genes and biological pathways that may underlie our identified GWAS findings. In genetic correlation and causal inference analyses, we delineated potentially causal links between common neurodegenerative and affective disorders and volumes of the mammillary bodies. Given that the mammillary bodies are a relatively understudied component of the Papez circuit and existing evidence points towards their involvement in different neurological and psychiatric disorders, our findings provide a unique and valuable foundation to understand their genetic determinants and thereby lay the groundwork for further delineation of their precise role in the pathogenesis of many brain disorders.

## ACKNOWLEDGEMENTS

We would like to thank the Rhineland Study team for supporting the data acquisition and management.

## Author contributions

MAI, MMBB, and NAA conceptualized the project. The first draft of the manuscript was written by MAI and NAA. Data were analyzed by MAI, ISR, DL and NAA. All authors provided technical, statistical, methodological advice and provided critical feedback and contributed to the writing and revision of the final version of the manuscript.

## Data availability

The data from the RS used in this manuscript is not publicly available due to data protection and privacy regulations. For the RS, access can be obtained by submitting a formal request to the Data Access Committee (RSDUAC) for academic, non-commercial research purposes, and in accordance with the RS’s Data Use and Access Policy (https://www.rheinland-studie.de/en/participation/ rules-of-use/). The committee will review all requests within 4 weeks. Access may be subject to a completed data transfer agreement. We used publicly available GWAS summary statistics for chronotype (https://journals.sagepub.com/doi/full/10.1177/0748730420935328), daytime napping and sleep duration (https://www.nature.com/articles/s41467-020-20585-3), cognition (https://www.nature.com/articles/s41588-018-0152-6), Alzheimer’s disease from Kunkle et al. (https://ftp.ebi.ac.uk/pub/databases/gwas/summary_statistics/GCST007001-GCST008000/GCST007511/), and Bellenguez et al. (https://www.nature.com/articles/s41588-022-01024-z), Parkinson’s disease (https://pmc.ncbi.nlm.nih.gov/articles/PMC8422160/), Major depressive disorder (https://figshare.com/articles/dataset/GWAS_summary_statistics_for_major_depression_PG <u>C_MDD2025_/27061255</u>) and Schizophrenia (https://figshare.com/articles/dataset/scz2022/19426775) and brain imaging-derived endophenotypes (https://www.nature.com/articles/s41588-019-0516-6). GWAS summary statistics for dementia, dementia including outpatient diagnoses (DEMENTIA_INCLAVO), psychological development disorders (PSYCHDEV), opioid-related disorders (OPIOIDS), habit and impulse disorders (HABIT), and severe stress-related disorders (STRESSOTH) were obtained from the FinnGen study and are publicly available through the FinnGen data repository (https://www.finngen.fi/en/access_results) and Google Cloud Storage (https://storage.googleapis.com/finngen-public-data-r13/summary_stats/). The specific FinnGen endpoints used in this study were F5_DEMENTIA, F5_DEMENTIA_INCLAVO, F5_PSYCHDEV, F5_OPIOIDS, F5_HABIT, and F5_STRESSOTH. GWAS summary statistics based on the Million Veteran Program (MVP) for migraine with aura (Phe_340_1) and abnormal involuntary movements (Phe_350_1) were obtained from https://ftp.ncbi.nlm.nih.gov/dbgap/studies/phs002453/phs002453.v1.p1/analyses/GIA/. GWAS summary statistics for memory impairment were obtained from https://ftp.ebi.ac.uk/pub/databases/gwas/summary_statistics/GCST90475001-GCST90476000/GCST90475544/.

---

**Supplementary Figure 1:**
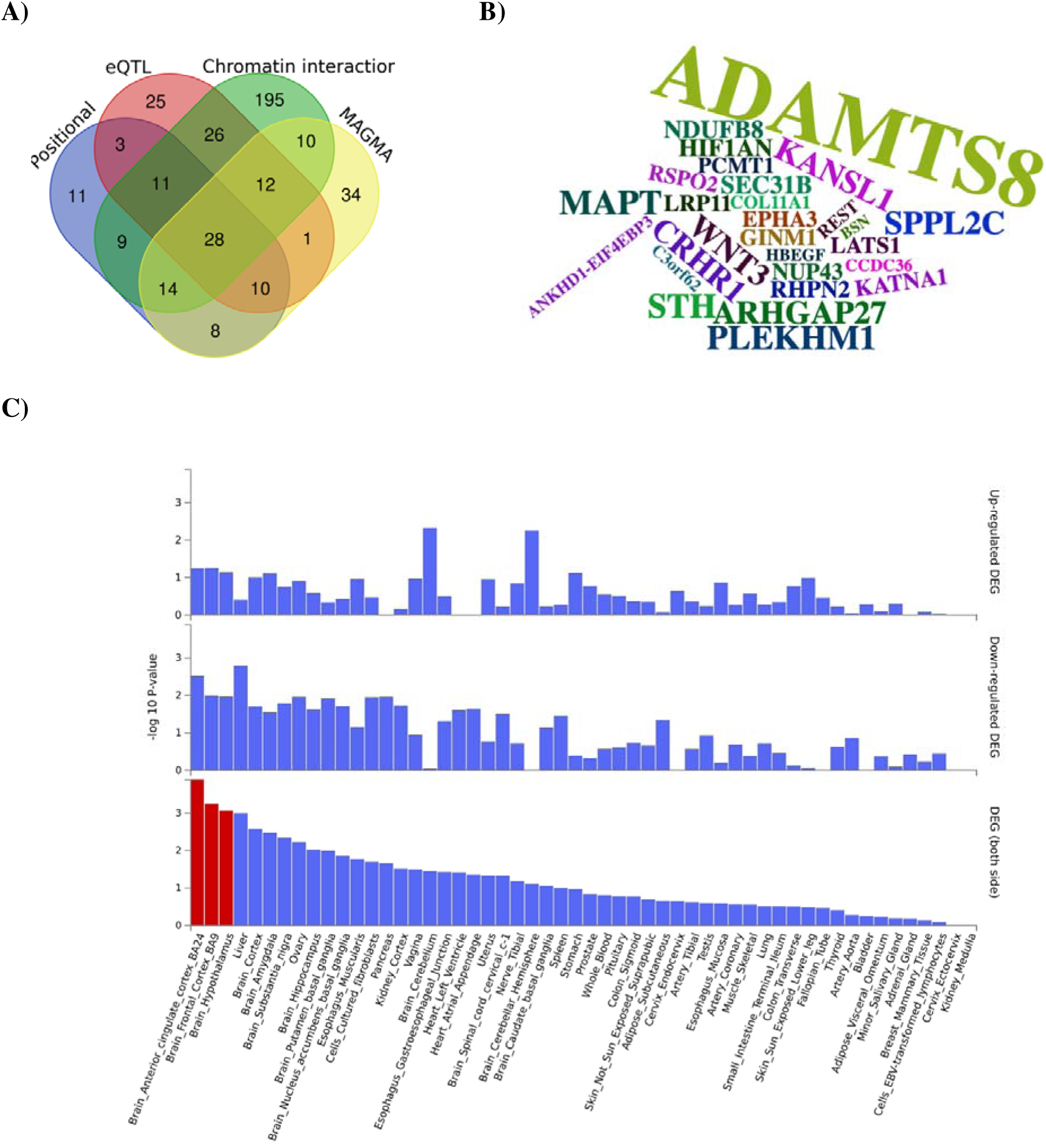
**A)** Gene mapping of the multivariate-GWAS results based on positional and functional annotation of lead (r^2^<0.1) and candidate SNPs (r^2^>0.6) via positional, expression quantitative trait loci (eQTL), chromatin interaction and MAGMA gene-based mapping. **B)** Word cloud displaying genes implicated by all four strategies, with word size proportional to the -log (P-value) values from the GWAS. **C)** GTEx v8 tissue-specific gene expression enrichment analysis of the 28 mapped genes across 53 tissue types using GENE2FUNC (FUMA). Bar plots display −log (P-value) for each tissue across three gene expression categories: up-regulated differentially expressed genes (DEGs, top), down-regulated DEGs (middle), and DEGs from both directions combined (bottom). Red bars indicate tissues surviving Bonferroni correction (P < 0.05); blue bars represent nominally enriched tissues.

**Supplementary Figure 2:**
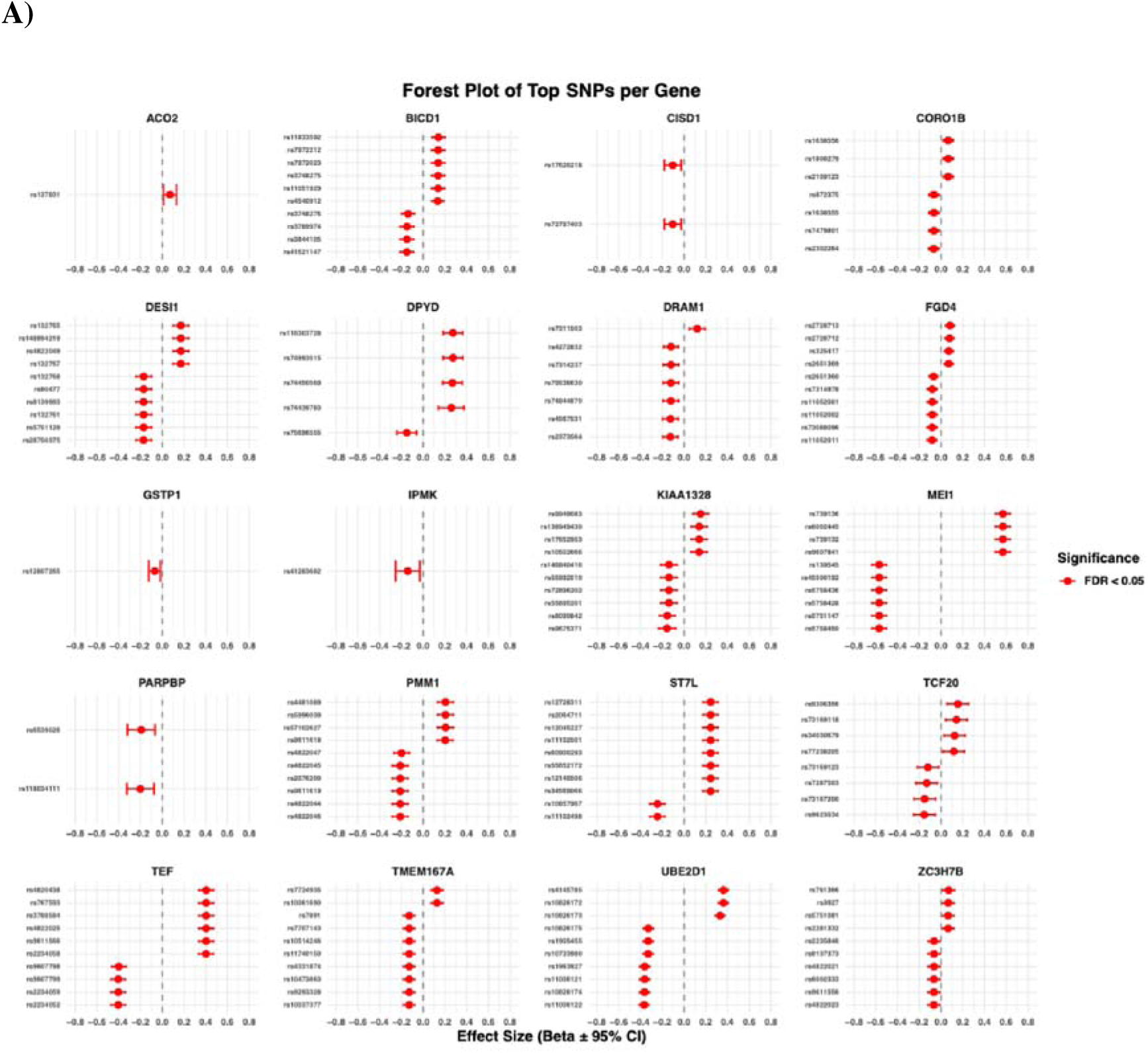
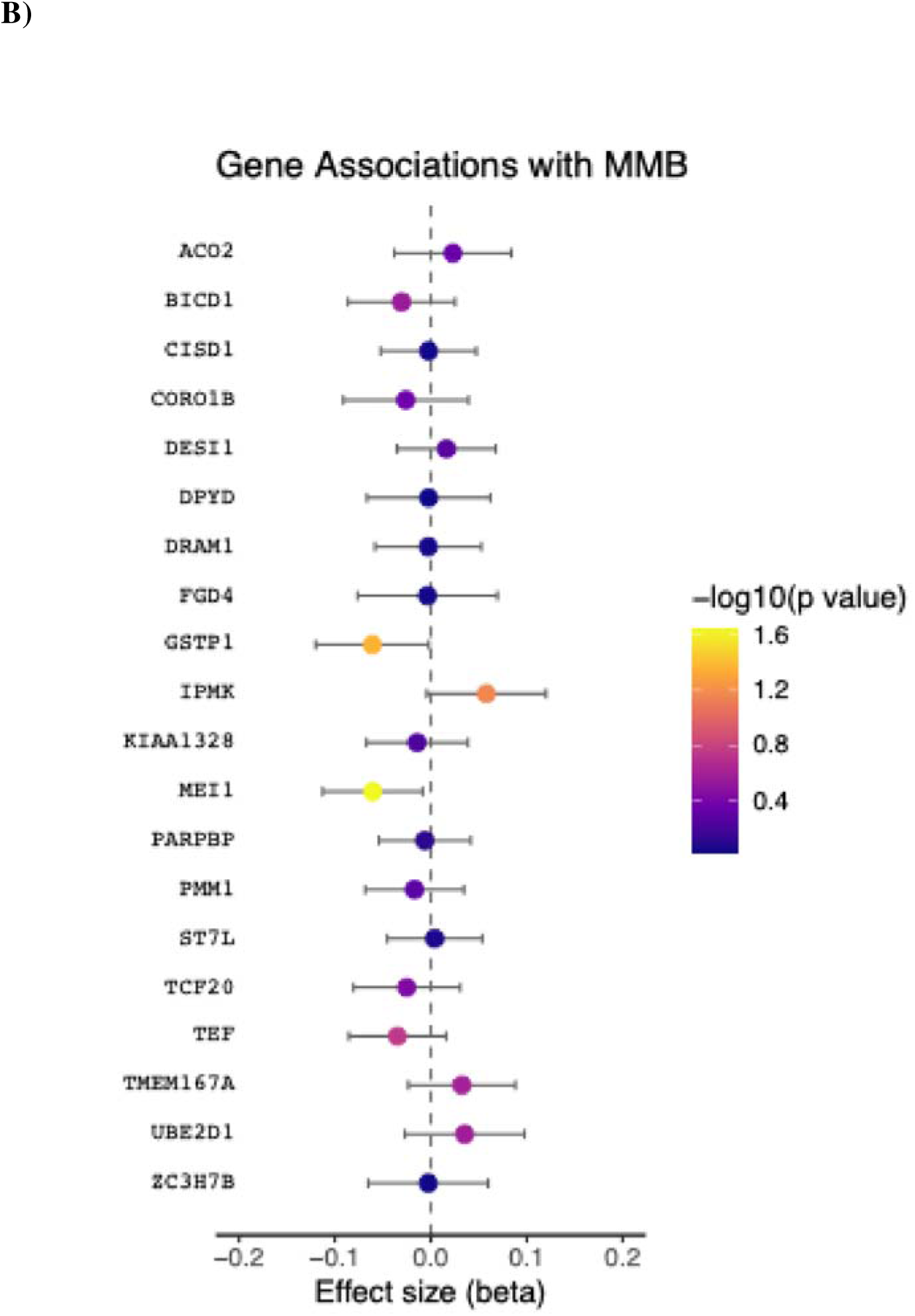
Transcriptomic analysis of candidate genes for mammillary body volume in the Rhineland Study. **A)** Expression quantitative trait loci (eQTL) associations of candidate genes based on whole-blood transcriptomics. **B)** Associations between whole-blood gene expression levels and mammillary body volume. Abbreviations: MMB = mammillary bodies

**Supplementary Figure 3:**
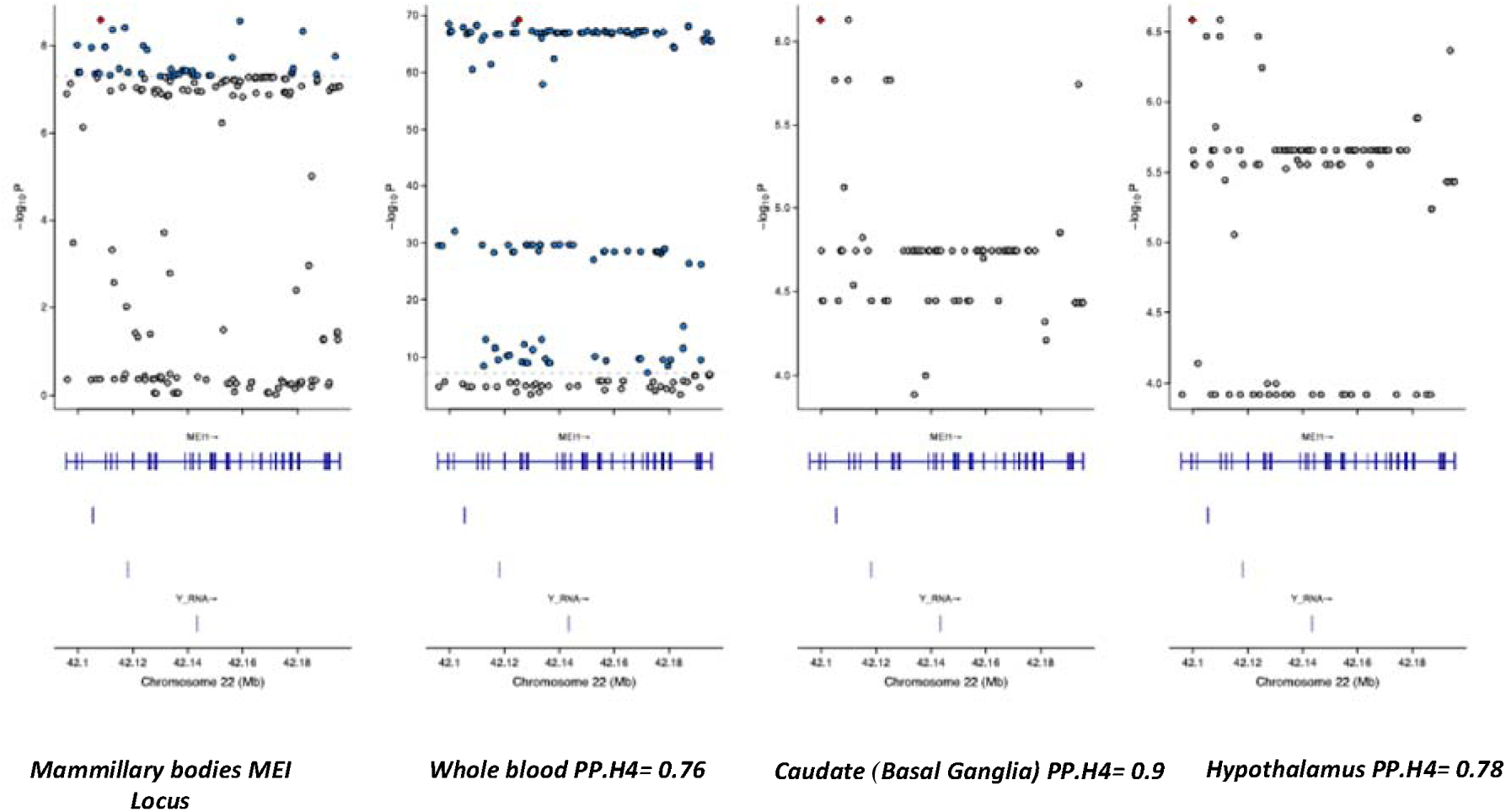
Colocalization of genetic variants associated with mammillary body volume and tissue eQTLs. Colocalization of genetic variants in the *MEI1* locus associated with mammillary body volume and eQTLs across whole blood and other brain tissues that surpassed a posterior probability threshold of 0.75 (i.e., PP.H4 > 0.75). PPH4 denotes the posterior probability hypothesis H4, corresponding to a shared causal variant between mammillary body volume and eQTL signals, indicating colocalization at the *MEI1* locus.

**Supplementary Figure 4:**
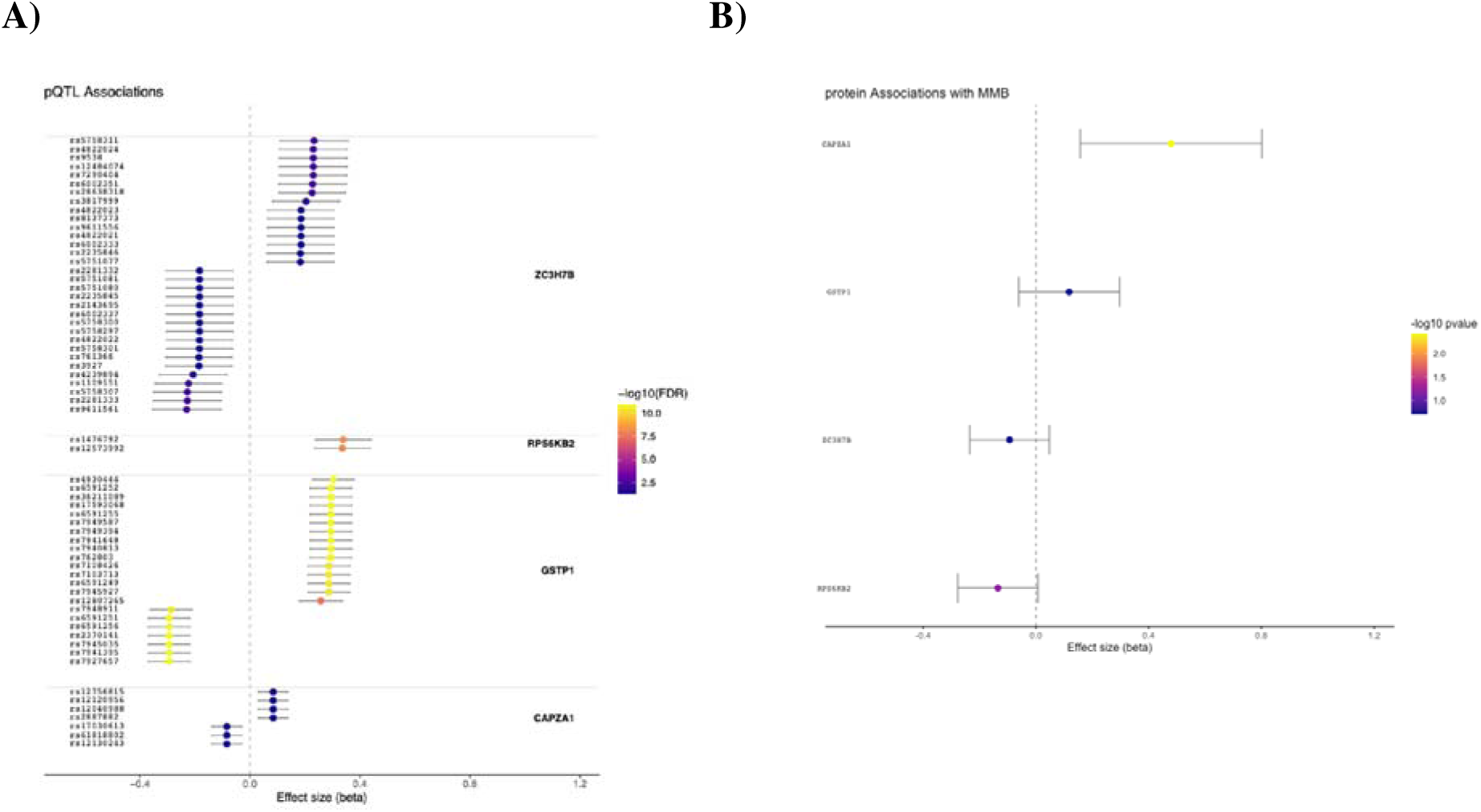
Proteomic analysis of candidate genes associated with mammillary body volume using the SEER platform in the Rhineland Study. **A)** Protein quantitative trait loci (pQTLs) associations in whole blood. **B)** Protein expression associations with mammillary body volume.

